# Effects of supervised high-intensity hardstyle kettlebell training on grip strength and health-related physical fitness in insufficiently active older adults: The BELL pragmatic controlled trial

**DOI:** 10.1101/2021.06.27.21259191

**Authors:** Neil J. Meigh, Justin W.L. Keogh, Ben Schram, Wayne Hing, Evelyne N. Rathbone

## Abstract

The Ballistic Exercise of the Lower Limb (BELL) trial examined efficacy and safety of a pragmatic hardstyle kettlebell training program in older adults. Insufficiently active men and women aged 59-79 years, were recruited to a 6-month repeated measures study, involving 3-months usual activity and 3-months progressive hardstyle kettlebell training. Health-related physical fitness outcomes included: grip strength [GS], 6-min walk distance [6MWD], resting heart rate [HR], stair-climb [SC], leg extensor strength [LES], hip extensor strength [HES], Sit-To-Stand [STS], vertical jump [CMVJ], five-times floor transfer [5xFT], 1RM deadlift, body composition (DXA), attendance, and adverse events. Sixteen males (68.8 ± 4.6 yrs, 176.2 ± 7.8 cm, 90.7 ± 11.0 kg, 29.2 ± 2.6 kg/m^2^) and sixteen females (68.6 ± 4.7 yrs, 163.9 ± 5.4 cm, 70.4 ± 12.7 kg, 26.3 ± 4.9 kg/m^2^) were recruited. Compliance to the supervised exercise program was very high (91.5%). Kettlebell training increased GS (R: MD = 7.1 kg 95% CI [4.9, 9.3], *p* < 0.001, L: MD = 6.3 kg 95% CI [4.1, 8.4], *p* < 0.001), 6MWD (41.7 m, 95% CI [17.9, 65.5], *p* < .001), 1RM (16.2 kg, 95% CI [2.4, 30.0], *p* = 0.013), 30s STS (3.3 reps, 95% CI [0.9, 5.7], *p* = 0.003), LES (R: MD = 61.6 N, 95% CI [4.4, 118.8], *p* = 0.028), HES (L: MD = 21.0 N, 95% CI [4.2, 37.8], *p* = 0.007), appendicular skeletal lean mass (MD = 0.65 kg, 95% CI [0.08, 1.22], *p* = 0.016), self-reported health change (17.1%, 95% CI [4.4, 29.8], *p* = 0.002) and decreased SC time (2.7 sec, 95% CI [0.2, 5.2], *p* = 0.025), 5xFT time (6.0 sec, 95% CI [2.2, 9.8], *p* < 0.001) and resting HR (7.4 bpm, 95% CI [0.7, 14.1], *p* = 0.032). There were four non-serious adverse events. Mean individual training load for group training sessions during the trial was 100,977 ± 9,050 kg. High-intensity hardstyle kettlebell training was well tolerated and improved grip strength and measures of health-related physical fitness in insufficiently active older adults.

## 1. Introduction

Aging is associated with a variety of biological changes that can contribute to decreases in skeletal muscle mass, strength, and function. Such losses decrease physiologic resilience and increase vulnerability to morbidity and mortality. Countering muscle disuse through structured exercise programs, particularly resistance training, is a powerful intervention to combat physiological vulnerability and its debilitating consequences on physical function, mobility, independence, chronic disease management, psychological well-being, and mental health ^(1)^. However, only 1 in 10 Australians over 50 years of age does enough exercise to gain any cardiovascular benefit, with estimates among the wider population of one in four people not being sufficiently active ^(2)^, with a similarly small proportion (9.6%) performing resistance training consistent with the guidelines ^(3)^. Solutions which slow the natural history of functional decline are needed to promote greater engagement in structured physical activity among older adults.

Hardstyle kettlebell training claims to improve measures of health-related physical fitness ^(4)^. Trials with younger participants report improvements in upper limb endurance ^(5)^, dynamic balance and vertical jump ^(6)^, leg strength and trunk endurance ^(7)^, standing long jump and grip strength ^(8)^, VO_2_ ^(9)^, and 1RM barbell deadlift ^(10)^. A profile of the kettlebell swing in novice older adults shows peak net ground reaction force is higher during a swing with an 8 kg kettlebell than a deadlift with 32 kg ^(11)^, highlighting the potential utility of the swing in exercise prescription. Anecdotally, older adults can and do engage in kettlebell training, but there is little data about the effects of kettlebell training in an older population.

Very few investigations of kettlebell training have involved older adults. Only two longitudinal trials have been conducted to address conditions associated with aging; sarcopenia in females ^(12)^ and Parkinson’s disease ^(13)^. These studies reported improvements in sarcopenia index, grip strength, back strength, respiratory function, and inflammatory markers ^(12)^, Timed Up and Go, upper limb strength, and lower limb strength ^(13)^. Although these results are encouraging, a focus on clinical conditions and limited reporting of trial protocols in these studies, offer little guidance for training otherwise healthy older adults. Furthermore, a recent review ^(14)^ identified no clinical practice guidelines or recommendations for using kettlebells with older adults. Most studies investigating the effects of kettlebell training are not representative of hardstyle techniques or training protocols. Investigation of the effects from highly controlled single kettlebell exercises with young healthy adults, provide policy makers with insufficient information about the safety or effectiveness of community-based group exercise programs for inactive men and women over 60 years of age, thus, a pragmatic assessment of typical training practices was required.

The aim of this study was to measure change in health-related physical fitness following 3-months of moderate- to high-intensity group hardstyle kettlebell training, in insufficiently active men and women over 60, in comparison with 3-months of usual activities of daily living. Additionally, exercise adherence and compliance rates, adverse events, and participant feedback about their experience were also collected to provide some information about the safety and feasibility of the exercise program. It was hypothesised that the intervention would be safe and effective for insufficiently active older adults to engage in group-based kettlebell training, with significant clinically meaningful improvements in measures of healthy aging compared to control. Our research question was: *can group-based hardstyle kettlebell training be used safely and effectively to engage* insufficiently active *older adults in the community and promote healthy aging?*

## 2. Material and methods

### 2.1. Study design and sample size

The BELL trial was an exploratory, single-centre, single cohort, repeated measures, controlled exercise intervention using hardstyle kettlebell training. Testing was conducted at baseline, week 4, week 13, week 19 and week 29. The study protocol had been published elsewhere https://osf.io/dz96p/. Scheduled test dates were changed due to the COVID-19 pandemic, which resulted in unequal periods between tests. Grip strength was selected as the primary outcome for clinical relevance, with measurements recommended in routine clinical practice and in community healthcare with older adults ^(15)^. Grip strength is a reliable surrogate for more complicated measures of arm and leg strength and a consistent predictor of falls and fractures in both sexes ^(16, 17)^. Low grip strength is a powerful predictor of functional limitations, poor health-related quality of life (HRQoL), strongly and inversely associated with all-cause mortality ^(18-20)^ and associated with higher annual healthcare costs ^(21)^. Secondary outcomes included a core set of clinical field tests of health-related physical fitness ^(22)^. Sample size was calculated based on the primary outcome from existing data ^(12)^ to detect an effect size change of 0.88 ^(23)^. A total of 19 participants were required to detect 95% power (1-*ß* = 0.05) and test the null hypothesis of equality (α = 0.05). Thirty-two participants were recruited to account for a 25% dropout and 15% for multivariable modelling. The current report comprises the findings and analysis of primary and secondary outcomes. A pragmatic approach was chosen to evaluate the effectiveness of the intervention in usual conditions, to identify variation between individuals, and maximise external validity ^(24, 25)^. Participants were not blinded to the hypothesis and resource constraints prevented blinding of data collection and analysis. A timeline for the study is presented in Figure 1.

**Fig. 1:**
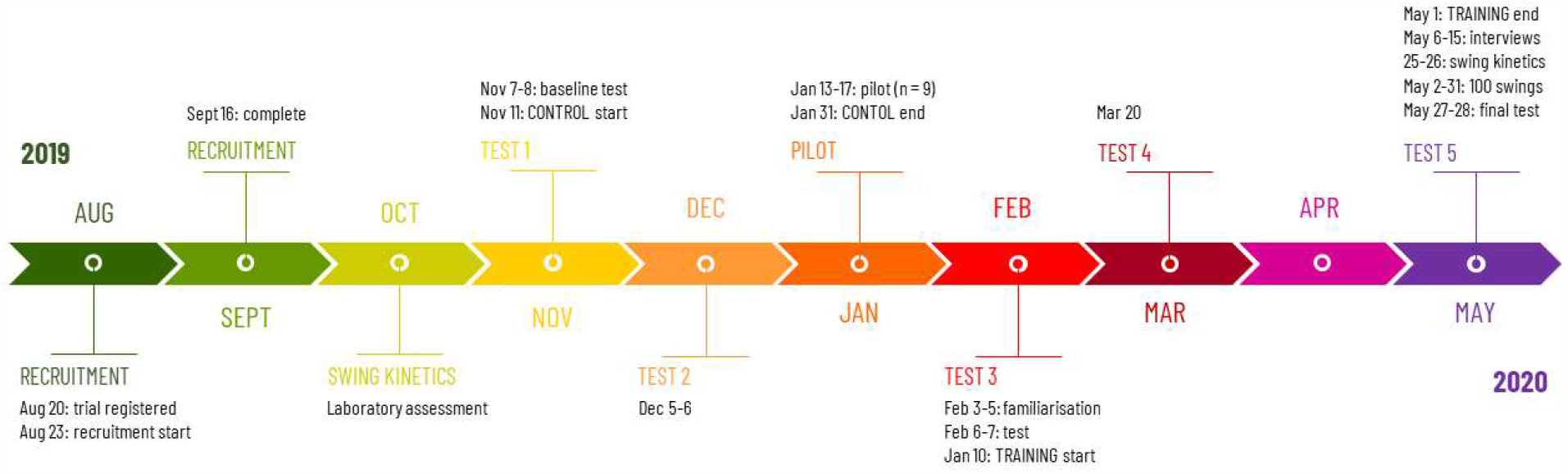
Study timeline

### 2.2. Ethical approval

All research activities were conducted in accordance with the *Declaration of Helsinki*. The trial was prospectively registered on the Australian New Zealand Clinical Trials Registry (ACTRN12619001177145) and approved by the Bond University Human Research Ethics Committee (BUHREC; Protocol number NM03279). Written informed consent was obtained by the lead investigator from all participants.

### 2.3. Participants

Independently living, apparently healthy but insufficiently active males and females were recruited via print media advertising from the central Gold Coast in south-east Queensland, Australia. A minor change was made to the inclusion criteria after trial registration, with the minimum age reduced to 59 years from the originally stipulated “60 years of age”. Exclusion criteria were as follows: medical conditions or medications known to affect musculoskeletal health which limit the capacity to perform moderate-to-high intensity exercise, recent surgery or trauma, uncontrolled cardiovascular or respiratory disease, engaged in a structured exercise program within the past nine months, malignancy, inability to perform a floor transfer independently, inability to comfortably lift the upper extremities overhead, unexplained pain with fundamental movements/activities e.g. sitting, walking, lifting, carrying, pushing, pulling or twisting, and presence of hazards which would prevent body composition analysis. Further exclusion was based on inability or unwillingness to attend three times weekly supervised training for the stipulated 3-month period. Participants completed a standardised pre-exercise screening tool and were asked to provide medical clearance from their General Practitioner. After commencement of the exercise period, it was noted that one participant was unable to perform a floor transfer unassisted, and one participant disclosed having osteoporosis and diabetic peripheral neuropathy. Participants would complete 12 weeks of usual activity (CON) and 12 weeks of kettlebell training (KBT).

### 2.4. Exercise intervention

Participants attended three-times weekly (Mon, Wed, Fri), 45-min, group classes at the Bond University High Performance Training Centre, Gold Coast, Australia, supervised by a Physiotherapist and certified (RKC) kettlebell Instructor. In addition, participants completed twice-weekly (Mon, Thur) prescribed home exercises. Program design was based upon the training principles and practices described by Tsatsouline ^(4)^ with exercises and delivery adjusted to meet individual limitations and the group dynamic, to maximise facilitators and reduce barriers to engagement. Classes were conducted face-to-face for six weeks, then remotely thereafter due to COVID-19 restrictions. The training period was preceded by a familiarisation week (2x 45-minute sessions) in which the participants were introduced to a standardised mobility drill (available as *supplementary data*) and the foundational kettlebell exercises of a swing, clean, military press, goblet squat and unloaded Turkish get-up. Kettlebell weights ranged from 4-80 kg. Participants were provided with a modified CR10 scale ^(26)^ for reporting sRPE. During the first two weeks, participants were advised to work at a relatively low intensity (2-4/10: “easy” to “somewhat hard”) with a low volume training load to minimise the likelihood of experiencing DOMS. From week three onward, participants were encouraged to work up to a sRPE of 5-7/10 (described as “hard” to “very hard”) as tolerated. Maximal effort (9-10/10) was discouraged. Where technique was acceptable and RPE appeared to be <4/10, participants were encouraged to increase exercise intensity (kettlebell weight). Exercises were modified where necessary to account for physical limitations or emergent biopsychosocial factors likely to effect performance. Home exercises, with an easy to moderate training load target (no upper limit), was performed at the participant’s discretion with an 8 kg kettlebell (provided). At the start of each group session, participants were asked to provide feedback regarding DOMS, adverse events, or health/medical conditions since the last training session.

All training sessions (Mon-Fri) commenced with the standardised mobility routine which served as a warm-up. Training sessions were planned based on a) physical capacity of the group, b) participant feedback, c) intent to offer variety, and d) plan to progress skill, intensity, and training load volume throughout the trial. Training plans were prepared within the preceding 36hrs. Participants were able to self-select weights and change any program variable within the group sessions. Participants completed a daily training record which included the exercise(s), number of sets and repetitions performed, and sRPE. Training records were collected at the end of each session then entered into a database for tracking and analysis. Due to COVID-19, ethics approval for face-to-face training was withdrawn, in effect from 23 March 2020. Training weeks 1-6 were delivered face-to-face; weeks 7-12 were delivered remotely, participants performing all remaining training independently at home Mon-Fri. Available kettlebells (8-40 kg) were given to participants to use at home from week 7, with each participant receiving two kettlebells in addition to the 8 kg provided at the start of the trial. These were roughly distributed based on physical capacity i.e., the stronger participants received heavier kettlebells. Reinstatement of ethics approval delayed final testing from 7-8 May to 27-28 May. In addition, all participants completed an international challenge to perform 100 kettlebell swings each day for the month of May, extending the planned training period by three weeks. Training material and programming was conceived, delivered, coordinated, and analysed by the lead investigator (NM). The full 3-month program is available as supplementary data.

### 2.5. Control activities

Participants were asked to continue their usual daily routines and refrain from taking up new structured exercise likely to influence the outcomes. In addition, participants were requested to not make significant dietary changes likely to alter body composition.

### 2.6. Outcomes

The planned schedule for data collection was changed due to the COVID-19 pandemic. The 4^th^ and 5^th^ rounds of testing, scheduled for weeks 22 (9-10 April) and 26 (7-8 May), took place in weeks 19 and 29 respectively, with some measures unable to be collected. Tests at weeks 0, 4, 13 and 19 were conducted at the Bond Institute of Health & Sport (*schedule of data collection available as supplementary data)*. Tests at weeks 0, 4 and 13 were conducted in a 5hr session, with participants completing a series of five 15 min stations, beginning with a fasted body composition scan (DXA). Tests in week 29 were completed in a single morning (*n* = 11). All DXA scans were conducted by the same licenced operator. Other data collection stations were conducted by a trained volunteer (or the lead investigator) following standard operating procedures prepared by the lead investigator. Tests involving prolonged close physical contact were omitted from the week 19 schedule due to COVID-19 restrictions. At week 29, only GS, 6MWD and 5xFT were collected for participants over 70 years, with testing conducted outdoors; an equidistant 6MWT was conducted on a straight, level, concrete walking track in a shaded area, and the 5xFT was conducted on level grass. All data, including DXA scans, were processed and analysed by the lead investigator (NM). Invalid data were deleted from one participant where outcome measures had been adversely affected by hip pain (5xFT, SC, 6MWT, STS, sit-and-reach, 1RM and CMVJ); the participant withdrew at the end of the control period and did not participate in the training. Invalid grip measures were deleted from week 0 and week 4 from one participant who had sustained a wrist injury and was unable to perform the test as required. A single resting systolic blood pressure reading >180mmHg was deleted from baseline data; the participant was subsequently treated by his GP and remained normotensive thereafter. Hip extension RFD could not be calculated at week 13 due to an error in the load cell rate of data capture.

#### 2.6.1. Grip strength

Grip strength was measured by Jamar handheld isometric dynamometer following a modified Southampton protocol ^(27)^.

#### 2.6.2. Anthropometrics

Age was obtained from self-report at baseline. Height was assessed without shoes using the stretch stature method with a wall-mounted stadiometer (Model Holtain 602VR, Seritex, New Jersey, USA). Weight in light clothing was measured using a calibrated scale (Model WM204, Wedderburn, Ingleburn, Australia). Body mass index (BMI) was calculated as mass divided by height squared.

#### 2.6.3. Cardiorespiratory endurance

Two tests of physical capacity were performed to examine cardiorespiratory capacity. Aerobic fitness was assessed using a 6-minute walk ^(22, 28)^ conducted according to American Thoracic Society guidelines ^(29)^, with 6MWD, HR, and RPE ^(30)^ recorded. Predicted distance (6MWD_pred_) was calculated using a validated equation ^(31)^.

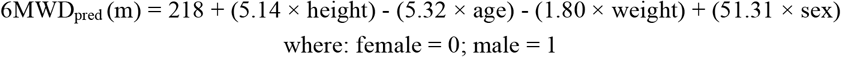

Submaximal aerobic capacity was measured using a stair climb test ^(32)^ with participants ascending a vertical displacement of 13.65m via seven flights of stairs having a 34° inclination. Four flights had 15 steps, 3 flights had 6 steps (totalling 78 steps), and there were approximately 21 steps on level ground between flights. Step height measured 17.5cm. Participants were instructed to climb the stairs as quickly as possible, with standardized verbal encouragement at each flight given by the examiner. Participants were asked to not use the handrail unless necessary and ensure one foot was in contact with the floor at all times i.e., walk not running. The time taken to climb the stairs and finish with both feet on the top step was designated the stair-climb time. The test was performed only once. Work performed (W) was calculated using the formula: W = *m* × *g* × *h*, where *m* is participant mass (kg), *g* is acceleration due to gravity (9.81m.s^-2^), and *h* is vertical displacement (metres). Power (P) was obtained by dividing work by stair-climb time. Age-predicted VO_2_max_pred_ ^(33)^ and estimated VO_2_max_est_ from SCT ^(32)^ were calculated using the formulae:

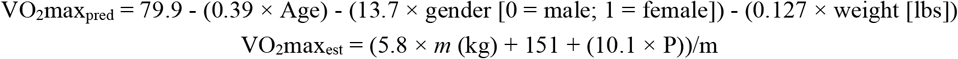

#### 2.6.4. Muscular strength and power

Maximal isometric leg extension ^(34)^ and hip extension ^(35, 36)^ were performed to examine lower limb strength. Force and RFD were recorded using a strain gauge (DBB Series S-Beam Load Cell, Applied Measurements, Berkshire, UK). Isometric leg extension was performed with the knee supported at 60° of flexion using a custom-made seat without back or arm rests (Fig. 2); participants sat upright with forearms crossed at the chest to perform the test. Hip extension was performed on a firm plinth (Fig. 3). Pelvic movement was restrained by a belt, the calcaneus suspended 8cm above the plinth with the participant’s forearms crossed at the chest to perform the test. *Relative* lower limb power was measured using the Sit To Stand App ^(37)^ using a height adjustable chair. A second seat of fixed dimensions was introduced at week 4 to improve standardisation of the tibia and femur in the start position, allowing a comparison of STS methods. Counter-movement vertical jump was performed as a functional expression of *absolute* lower limb power ^(38)^, with jump height determined from a floor-mounted force flatform (AMTI, Watertown, NY, USA) by applying the Impulse-momentum (IM) relation to the force-time curve ^(39)^.

**Fig. 2.**
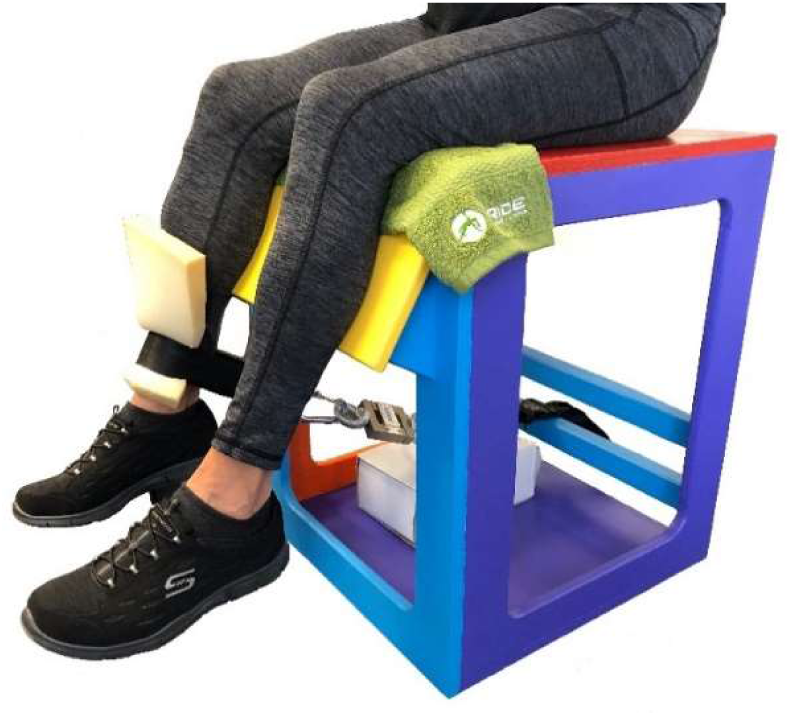
Leg extension strength test.

**Fig. 3.**
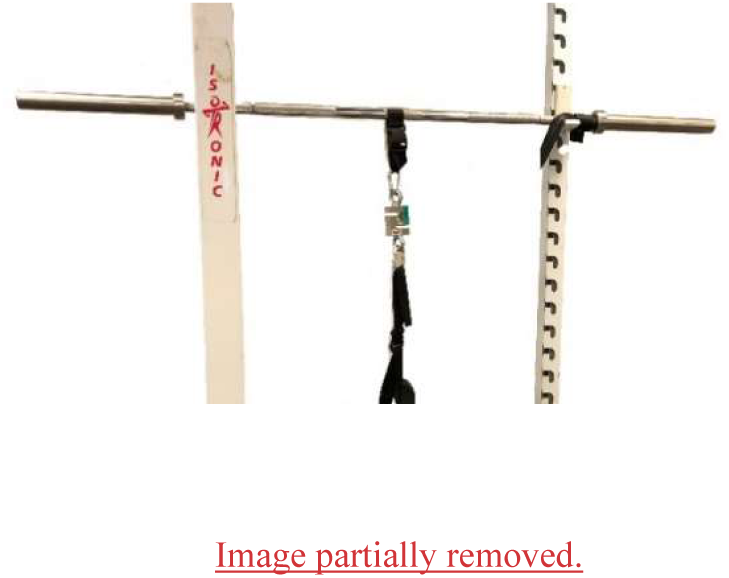
Hip extension strength test.

#### 2.6.5. Muscular endurance

Lower limb muscular endurance was examined using the 30s sit-to-stand test following a standardised procedure. ^(22, 40, 41)^.

#### 2.6.6. Flexibility

Flexibility was examined by fingertip-to-floor test ^(42, 43)^. A sit-and-reach test (Figure Finder Flex Tester, Novel Products, Rockton, USA) was also introduced at week 4 to limit movement at the knee ^(44)^.

#### 2.6.7. Body composition

A DXA scan (General Electric, GE, Lunar Prodigy, Madison, WI, USA) was conducted for each participant to determine body composition (fat and lean mass). The scanner was calibrated each morning using a manufacturer’s “phantom” following quality assurance and quality control procedures. Participants were required to wear only light clothing with metal objects removed, and positioned according to the Nana protocol ^(45)^. Results were analysed using the commercial software provided with the machine (enCORE software, version 17, GE, Lunar Prodigy, Madison, WI, USA) using regions of interest (ROI) recommended by the International Society for Clinical Densitometry official position ^(46)^. All scans were performed by the same technician, with intra-rater reliability and precision of DXA in evaluating BC previously described ^(47, 48)^. All ROI analysis was performed by the lead investigator (NM). Appendicular skeletal muscle mass (ASM) was calculated as the sum of the muscle mass of the four limbs. Appendicular skeletal muscle mass (also described as the Skeletal Muscle Mass Index (SMI) ^(49)^) was adjusted for height, calculated as ASM/height^2 (15)^. Body composition was also measured by BIA (Tanita MC-980MA PLUS, Tokyo, Japan) following a procedure previously described ^(50)^. The cross-validated Sergi equation ^(51)^ was used to calculate ASM from BIA data.

#### 2.6.8. Functional capacity

Functional capacity was examined using a 5-times floor transfer test ^(52, 53)^, predicted 1RM kettlebell deadlift using the two-point method ^(54)^, and excursion of centre of pressure during quiet standing balance on a floor-mounted force flatform (AMTI, Watertown, NY, USA), eyes open and eyes closed ^(55, 56)^.

#### 2.6.9. Quality of life and sense of coherence

Health Related Quality of Life (HRQoL) and Sense of Coherence were examined using the self-administered SF-36 questionnaire ^(57)^ and 29-item SoC scale ^(58)^.

#### 2.6.10. Training load

Exercises and V-TL were not set a-priori as participant’s physical capacity was unknown to the investigator. All training (Mon-Fri) commenced with a standardised mobility routine which was used as a warm-up. At the end of each group training session, participants submitted a training record of the exercises performed, weights used, number of sets and repetitions completed, and sRPE. During intervention weeks 1-6, a paper record was collected by the lead investigator at the end of each session and transcribed to a database for analysis. During intervention weeks 7-12, daily training records (Mon-Fri) were submitted by each participant via Survey Monkey. Session V-TL (kg) was programmed at an individual level during the final two weeks of the training, to enable each participant to achieve a personal best (sessional training load volume) on the final day of the program.

#### 2.6.11. Attendance, compliance, and adverse events

Exercise adherence was recorded, with 100% attendance defined as completion of 36, ‘group’ sessions over the 3-month trial period. Data were collected, entered into a database, and analysed by the lead investigator. Compliance to prescribed home-exercise was also reported. On Wednesday and Friday sessions during intervention weeks 1-6, participants reported whether the prescribed home-exercise had been completed the previous day with a Y/N response. Attendance and compliance were recorded for 12 weeks, with 100% attendance and compliance defined as 60 training sessions (group and individual) over the 3-month trial period. To maximise attendance and compliance, participants received frequent individual and group encouragement, both publicly and privately. Recognition was given to overcoming challenges, extraordinary effort, and achieving a ‘personal best’. Training and communication promoted *group* engagement to foster a spirit of support, camaraderie, and healthy competition. A private Facebook page was heavily used to provide encouragement, maintain accountability, and foster a community spirit. Participants were encouraged to make use of the on-site coffee-shop after training and provided a limited number of drink vouchers. Adverse events were defined as any undesirable outcomes that may be related to the intervention, which were recorded by the lead investigator for analysis. At enrolment, participants were provided with an information sheet about DOMS. Participants were asked to report to the principal investigator any new, unusual, or worrying physical symptoms, with participants frequently asked about their health and wellbeing before session training commenced.

#### 2.6.12. Concomitant care

Advice and guidance were provided to those who experienced muscle soreness and adverse training-related symptoms.

### 2.7. Statistical analysis

Statistical analyses were undertaken using SPSS statistical software version 26.0 (SPSS Inc., USA). Descriptive statistics, expressed as mean (SD) for normally distributed continuous variables, were generated for participant characteristics and all dependent variables. Normality was checked through a combination of histograms, normal Q-Q plots and the Shapiro-Wilk test. Categorical variables were summarised using frequencies and percentages. Linear mixed models (LMMs) were applied to the 32 participants regardless of withdrawal or attendance, measured at 5 time-points to model the change in quantitative outcomes after adjusting for potential confounders which included age, sex, and previous training history. Time was treated as a fixed factor to enable the assessment of any statistically significant changes between specific time-points. The individual was treated as a random effect. Random intercepts and random slopes were investigated to determine the most suitable models. The final models were fitted with random intercept only, using the restricted maximum likelihood estimation method (REML). Residual diagnostics were used to check distributional assumptions. Effect sizes (ES) were calculated and interpreted using Lenhard ^(59)^ and Magnusson ^(60)^, quantified as trivial < 0.20, small 0.20-0.59, moderate 0.60-1.19, large 1.20-1.99, very large 2.0-3.99, and extremely large ≥ 4.0 ^(61)^. Statistical significance was set at the 0.05 level *a priori*.

## 3. Results

### 3.1. Participant characteristics at baseline

Participant flow through the study is presented in Fig. 4. In total, 32 eligible participants completed all outcome measures at baseline. Three participants withdrew during the control period: medical condition (*n* = 1), injury (*n* = 1) and no longer able to attend (*n* = 1). Twenty-nine participants commenced the intervention. Five participants withdrew during the intervention: unexpected work (*n* = 1), substance abuse and mental health (*n* = 1), back pain (*n* = 1), viral infection (*n* = 1), uncontrolled hypertension: GP requested (*n* = 1). Participant characteristics at baseline are presented in Table 1. At baseline, males were significantly taller and heavier than females.

**Fig. 4.**
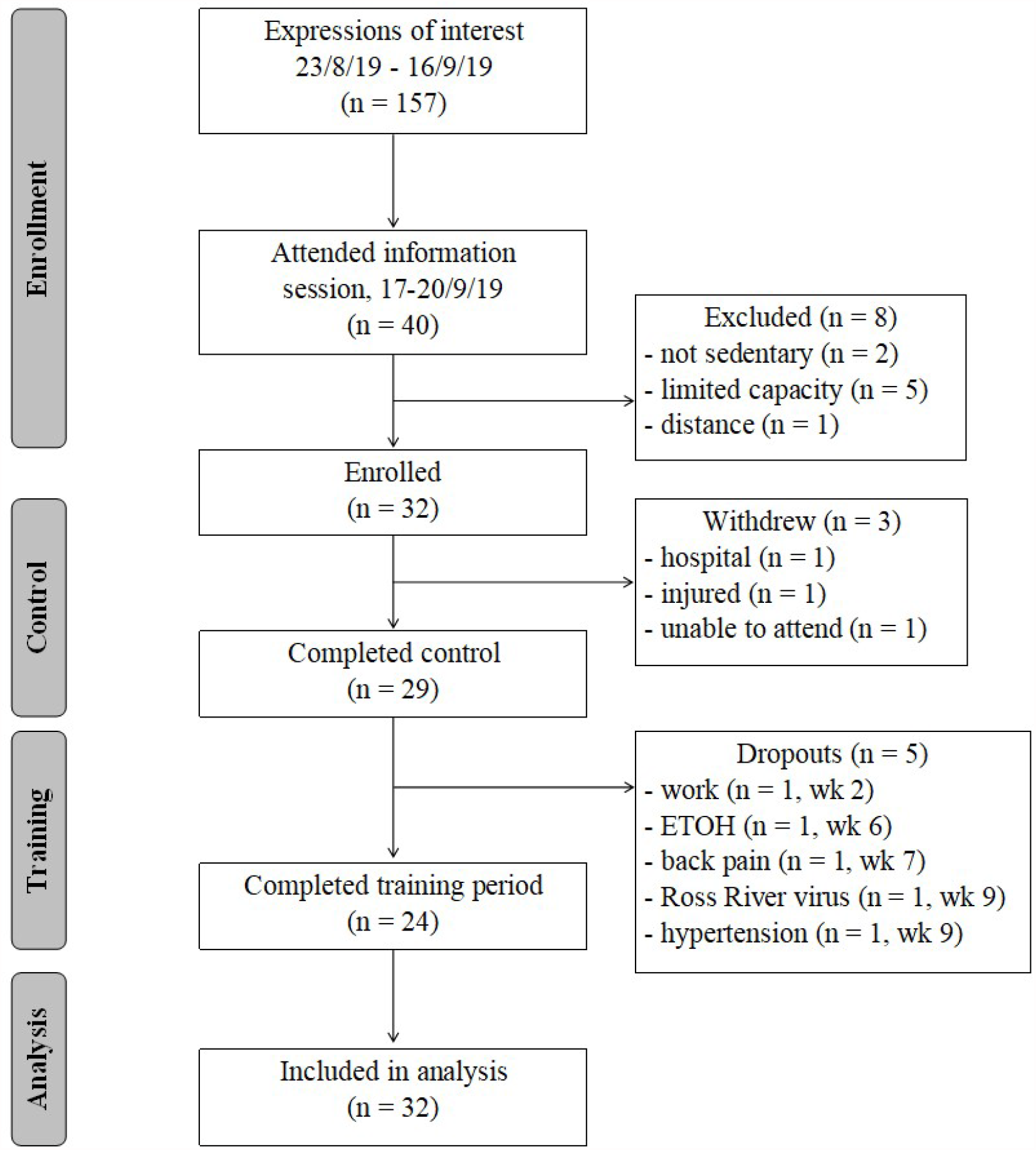
CONSORT diagram of participant flow through the BELL trial study. Abbreviation: ETOH, alchol

**Table 1.** Participant characteristics at baseline

| Characteristic | Males ($n = 16$) | Females ($n = 16$) |
| --- | --- | --- |
| Age (years) | 68.8 (4.6) | 68.6 (4.7) |
| Height (cm) | 176.2 (7.8) | 163.9 (5.4) |
| Weight (kg) | 90.7 (11.0) | 70.4 (12.7) |
| BMI (kg/m<sup>2</sup>) | 29.2 (2.6) | 26.3 (4.9) |

*Data are presented as mean (SD).*

Sixteen-week change in outcomes from mixed effects modelling are presented in Table 2. Pairwise comparison pre- to post-training (week 13 to week 29) are presented in Table 3. There was a large (δ > 1.4) pre- to post-training change in grip strength of 7.1 kg (95% CI [4.9, 9.3], *p* < 0.001) in the right hand, and 6.3 kg (95% CI [4.1, 8.4], *p* < 0.001) in the left hand, exceeding the minimum clinically important difference of 5.0 - 6.5 kg ^(62)^.

**Table 2.** Estimated regression coefficients from mixed effects modelling to show the effect of training over time (*N* = 32).

| Outcome |  | Baseline (95% CI) | Difference (95% CI) at |  |  |
| --- | --- | --- | --- | --- | --- |
|  |  |  | 13 weeks | 19 weeks | 29 weeks |
| Grip strength (kg) | *right* | 29.1 (25.3, 32.8) | 0.6 (-0.9, 2.1) | 4.2 (2.5, 5.8)*** | 7.8 (6.0, 9.5)*** |
|  | *left* | 27.9 (24.4, 31.3) | 0.3 (-1.2, 1.8) | 4.0 (2.4, 5.6)*** | 6.6 (4.9, 8.2)*** |
| 6-min walk distance (m) |  | 599.8 (577.4, 622.3) | 30.2 (14.0, 46.9)*** |  | 71.6 (53.9, 89.3)*** |
| Stair climb time (sec) |  | 48.1 (44.2, 52.0) | -3.6 (-4.7, -2.4)*** | -5.4 (-6.6, -4.1)*** | -6.2 (-8.0, -4.5)*** |
| Systolic blood pressure (mmHg) |  | 124.5 (118.8, 130.2) | -5.9 (-12.2, 0.4) |  | 6.9 (-1.9, 15.7) |
| Resting heart rate (bpm) |  | 69.4 (64.5, 74.3) | 10.1 (5.4, 14.8)*** |  | -7.4 (-14.1, -0.7)* |
| Knee extension peak force (N) | *right* | 321.6 (288.3, 355.0) | -9.9 (-38.5, 18.8) |  | 51.7 (9.4, 94.1)** |
|  | *left* | 285.9 (254.3, 317.4) | 12.4 (-11.4, 36.3) |  | 62.2 (26.3, 98.1)*** |
| Knee extension RFD (N.s<sup>-1</sup>) | *right* | 133.5 (118.0, 148.9) | 46.2 (-104.1, 196.6) |  | -114.2 (-334.4, 105.9) |
|  | *left* | 584.3 (455.9, 712.7) | 268.4 (104.8, 432.0)** |  | 61.4 (-173.6, 296.4) |
| Knee extension peak torque (N.m) | *right* | 133.5 (118.0, 148.9) | -4.2 (-16.1, 7.6) |  | 20.7 (3.1, 38.3)** |
|  | *left* | 118.7 (104.1, 133.3) | 4.8 (-4.8, 14.4) |  | 25.0 (10.3, 39.7)*** |
| Hip extension peak force (N) | *right* | 160.1 (146.4, 173.8) | 6.9 (-0.3, 14.1) |  | 17.7 (6.9, 28.4)** |
|  | *left* | 164.7 (150.0, 179.4) | 7.7 (-0.8, 16.1) |  | 28.7 (16.3, 41.0)*** |
| Hip extension RFD (N.s<sup>-1</sup>) | *right* | 240.1 (172.9, 307.3) |  |  | 156.6 (32.7, 280.6)* |
|  | *left* | 271.1 (167.2, 375.1) |  |  | 58.0 (-146.5, 262.4) |
| Hip extension peak torque (N.m) | *right* | 135.6 (122.5, 148.8) | 5.8 (-0.4, 11.9) |  | 14.2 (5.0, 23.4)** |
|  | *left* | 139.4 (125.9, 153.0) | 5.9 (-1.3, 13.0) |  | 22.1 (11.5, 32.7)*** |
| 30s Sit to Stand (W.kg<sup>-1</sup>) |  | 7.20 (6.86, 7.53) | 0.36 (0.03, 0.69)* |  | 0.48 (0.03, 0.94)* |
| Vertical jump (cm) |  | 13.5 (11.8, 15.2) | 0.0 (-0.6, 0.6) | 0.4 (-0.4, 1.3) | -0.2 (-1.3, 0.8) |
| 30s Sit to Stand (reps) |  | 14.2 (12.9, 15.4) | 1.7 (0.4, 2.9)* |  | 4.9 (3.1, 6.7)*** |
| Fingertip to floor (cm) |  | -2.7 (-6.7, 1.3) | -1.6 (-3.8, 0.7) |  | 2.5 (-0.8, 5.8) |
| Fat mass (kg) DXA |  | 30.92 (27.88, 33.96) | 0.02 (-0.42, 0.46) |  | -0.10 (-0.77, 0.58) |
| Lean mass (kg) DXA |  | 47.08 (43.38, 50.78) | 0.28 (-0.11, 0.66) |  | 0.99 (0.40, 1.57)*** |
| ASM (kg) DXA – *upper + lower limbs* |  | 21.34 (19.52, 23.16) | -0.08 (-0.35, 0.20) |  | 0.58 (0.17, 0.99)** |
| SMI (kg/m<sup>2</sup>) DXA |  | 7.27 (6.84, 7.70) | -0.01 (-0.11, 0.08) |  | 0.21 (0.07, 0.35)** |
| ASM (kg) BIA – *Sergi eqn* |  | 43.9 (40.3, 47.5) | 0.4 (-0.4, 1.2) |  | 0.7 (-0.4, 1.9) |
| 5x floor transfer time (s) |  | 41.8 (38.1, 45.5) | -5.9 (-8.4, -3.3)*** | -12.4 (-15.4, -9.3)*** | -11.8 (-15.3, -8.3)*** |
| 1RM (kg) – *predicted* |  | 69.5 (60.0, 78.9) | 5.6 (-1.3, 12.6) |  | 21.8 (11.6, 32.0)*** |
| Standing balance (cm<sup>2</sup>) – *eyes closed* |  | 23.3 (17.4, 29.1) | 2.1 (-3.5, 7.8) |  | 4.4 (0.4, 8.5)* |
| Health change - *SF36* |  | 52.4 (45.6, 59.2) | -0.1 (-8.5, 8.3) | 17.3 (8.2, 26.4)*** | 17.0 (7.8, 26.3)*** |
| Sense of coherence |  | 154.0 (144.7, 163.3) | -3.0 (-11.0, 5.0) |  | 2.6 (-8.3, 13.4) |

\* $p < 0.05$ \*\* $p < 0.01$ \*\*\* $p < 0.001$ significantly different to baseline.

**Table 3.** Linear mixed modelling results showing the mean differences between pre- and post-training after adjusting for age, sex and previous training history.

| Variable | | MD | SE | 95% CI | p-value | $\delta^a$ | % Change |
| --- | --- | --- | --- | --- | --- | --- | --- |
| Grip strength (kg) | *right* | 7.1 | 0.8 | (4.9, 9.3) | < 0.001 | 1.66 | 24.6 |
|  | *left* | 6.3 | 0.7 | (4.1, 8.4) | < 0.001 | 1.48 | 22.4 |
| Stair-climb time (s) |  | -2.7 | 0.9 | (-5.2, -0.2) | 0.025 | 0.55 | -5.6 |
| Stair-climb est. $\text{VO}_2$ ($\text{ml.kg}^{-1}.\text{min}^{-1}$) | | 2.3 | 0.7 | (0.2, 4.4) | 0.026 | 0.55 | 6.2 |
| 5x floor transfer (s) |  | -6.0 | 1.3 | (-9.8, -2.2) | < 0.001 | 0.80 | -14.3 |
| 6-min walk distance (m) |  | 41.7 | 8.7 | (17.9, 65.5) | < 0.001 | 0.85 | 7.0 |
| 30s Sit to Stand (reps) |  | 3.3 | 0.9 | (0.9, 5.7) | 0.003 | 0.66 | 23.0 |
| Predicted 1RM (kg) |  | 16.2 | 5.1 | (2.4, 30.0) | 0.013 | 0.57 | 23.3 |
| Knee extension – *peak force* (N) | *right* | 61.6 | 21.0 | (4.4, 118.8) | 0.028 | 0.52 | 19.1 |
|  | *left* | 49.8 | 18.7 | (-1.2, 100.7) | 0.059 | 0.47 | 17.4 |
| Hip extension – *peak force* (N) | *right* | 10.8 | 5.0 | (-3.0, 24.5) | 0.220 | 0.38 | 6.7 |
|  | *left* | 21.0 | 6.2 | (4.2, 37.8) | 0.007 | 0.60 | 12.7 |
| SF36 - *health change* |  | 17.1 | 4.4 | (4.4, 29.8) | 0.002 | 0.68 | 32.6 |
| DXA – *ASM* (kg) |  | 0.65 | 0.21 | (0.08, 1.22) | 0.016 | 0.55 | 3.1 |
| DXA – *sarcopenia index* ($\text{kg/m}^2$) | | 0.23 | 0.07 | (0.03, 0.42) | 0.012 | 0.57 | 3.1 |

Only outcome measures with a significant pre- to post-training change are displayed.
<sup>a</sup> effect size: week 13 to week 29 (pre- to post-training)

### 3.2. Adherence and compliance

Attendance rate was 91.5%. Twenty-one participants were absent from 81 of 956 potential Mon/Wed/Fri group training sessions. Eight participants had 100% group session attendance. The reasons for absence were: viral infection (*n* = 17), hospital/medical (*n* = 12), not disclosed (*n* = 7), unwell (*n* = 7), other infection (*n* = 6), muscle strain (*n* = 5), DOMS (*n* = 5), ‘life admin’ (*n* = 3), low back pain (*n* = 3), COVID-19 (*not infected*) (*n* = 3), bereavement (*n* = 2), mental health (*n* = 2), weather/transport (*n* = 2). Compliance with home-exercise was 88.7%. Seventy-one of 639 home sessions were reported as not completed. During intervention weeks 7-12, there were 35 recordings of training voluntarily undertaken on a weekend.

### 3.3. Change in secondary outcomes

#### 3.3.1. Cardiovascular endurance

Estimates of fixed effects showed a small (δ = 0.39) but significant reduction in resting HR from baseline of 7.4 bpm, but no significant change in systolic or diastolic blood pressure at any time points. At baseline, there was no significant difference in mean 6WMD (598.5 m) and age-predicted maximum (602.7 m). There was a significant moderate (δ = 0.85) 7% pre- to post-training increase in 6MWD of 41.7 m. There were statistically significant reductions in stair climb time at weeks 13, 19 and 29. Pairwise comparison pre- to post-training revealed a small (δ = 0.55) reduction of 2.5 seconds. At baseline, estimated VO2 calculated from stair climb time suggested a mean VO2 of 37.7 ml.kg^-1^.min^-1^ however, this was 54.2% higher than an age-predicted VO2 of 24.5 ml.kg^-1^.min^-1^.

#### 3.3.2. Muscular strength, power, and endurance

There were small (δ = 0.47 and 0.52) increases in knee extension peak force pre- to post-training, of 49.8 N and61.6 N in the left and right legs respectively, with no significant change in RFD. There were small to moderate changes (δ = 0.38 and 0.60) in hip extension peak force of 10.8 N and 21.0 N in the right and left hips, respectively, with no significant change in RFD. There was no significant change in lower limb power or vertical jump height however, there was a significant moderate (δ = 0.66) 23% increase of 3.3 repetitions performed during the 30sSTS test pre- to post-training.

#### 3.3.3. Flexibility

There was no significant change in flexibility at any time point.

#### 3.3.4. Body composition

DXA-derived appendicular lean mass significantly increased pre- to post-training by 0.65 kg (95% CI [0.08, 1.22], *p* = 0.016), with a corresponding increase in SMI of 0.23 kg/m^2^ (95% CI [0.03, 0.42], *p* = 0.012). There was no significant change in fat mass measured by DXA at any time point, and no significant change in muscle mass or fat mass measured by BIA between baseline and week 29.

#### 3.3.5. Functional capacity

Pre- to post-training, there was a significant moderate (δ = 0.8) reduction in 5-times floor transfer time of 6.0 seconds, (95% CI [9.8, 2.2], *p* < 0.001) and a significant 23.3% increase (δ = 0.57) in predicted 1RM of 16.2 kg (95% CI [2.4, 30.0], *p* = 0.013). There was no significant change in quiet standing balance.

#### 3.3.6. Quality of life & sense of coherence

There was a statistically significant 17% increase in the ‘overall health’ domain of the SF-36, but no significant change in any one sub-domain of health status, and no significant change in sense of coherence.

#### 3.3.7. Training load

Change in training load volume (kg and AUs) over time is presented in Fig. 5 Cumulative total training load volume for group sessions (Mon/Wed/Fri) was 1,022,220 kg for weeks 1-6 and 2,567,834 kg at week 12. Training load volume increased by 51.2% following the transition to home-only training. At 6 weeks (*n* = 28), mean external and internal training load was 36,067 (12,843) kg (range = 16,820 to 69,444 kg), and 3,430 (926) AUs (range = 2,115 to 5850 AUs), respectively. At 12 weeks (*n* = 24), mean training load was 100,914 (42,449) kg (range = 44,369 to 243,524) and 9,094 (1,727) AUs. External training load volume on the final day of the program was 29% higher than programmed: actual = 197,520 kg, mean = 8,230 (3623), range = 3,200 to 21,120 kg vs programmed = 153,070 kg, mean = 6,123 (2,281), range = 2,810 to 12,510 kg.

**Fig. 5.**
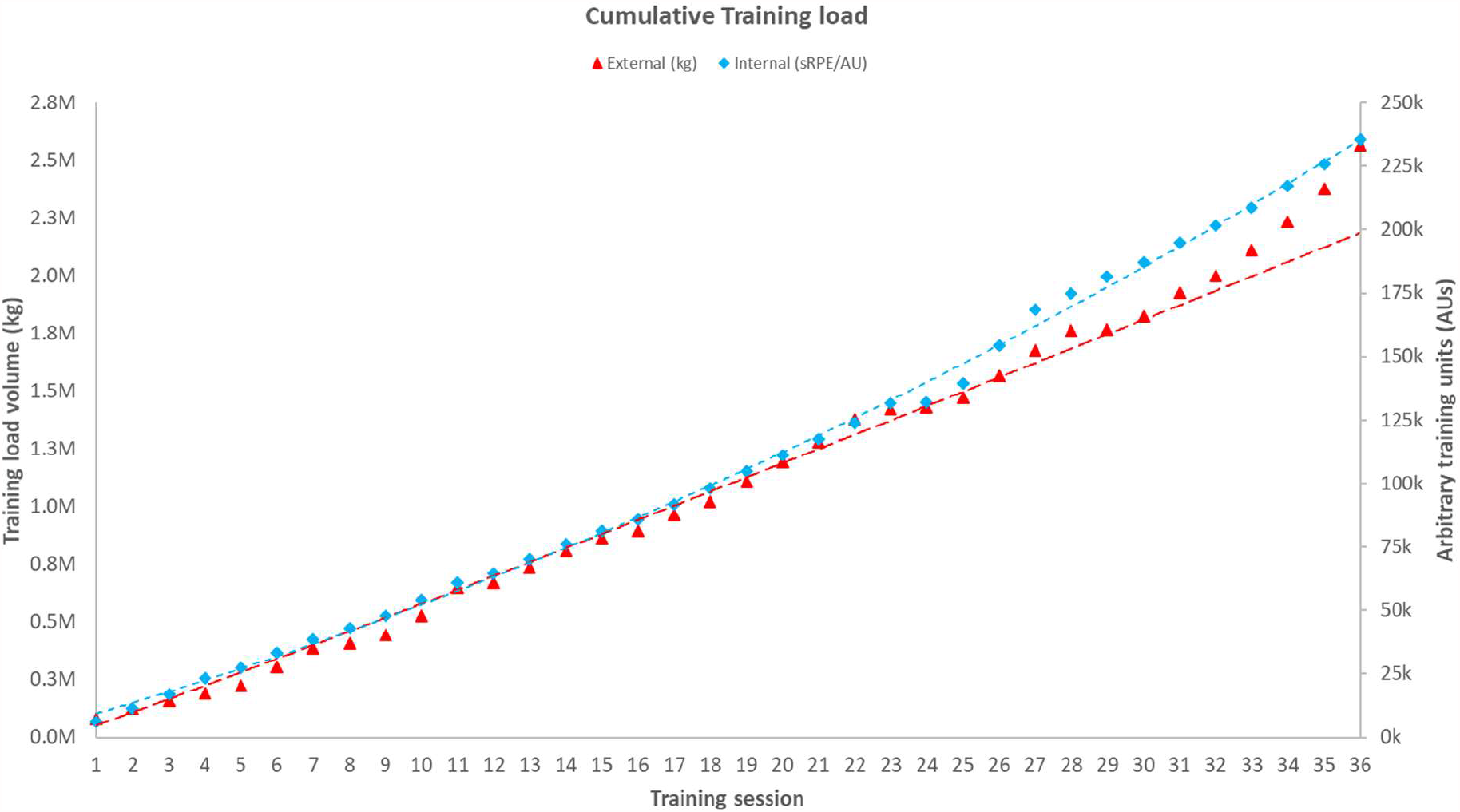
Cumulative group training load over time: kg (external training load) and arbitrary training unit (internal training load).

#### 3.3.8. Adverse events

As a normal and expected response, DOMS was not considered an adverse event however, 20/28 participants (71%) reported some degree of DOMS on at least one occasion, reported as mild (*n* = 5, 25%), moderate (*n* = 12, 60%) or severe (*n* = 3, 15%). All participants reported that their symptoms had resolved in ≤ 3 days, and 90% (*n* = 18) recorded that they were unconcerned by it. Four participants missed at least one training session due to muscle soreness. The number of participants experiencing DOMS was high but unsurprising and anticipated given the volume and intensity of the training. There were four non-serious adverse events. One female with a 50-year history of low back pain, experienced an exacerbation of symptoms following the final session in week 2. The participant completed the session without symptoms (no indication of mechanical stress/strain), which were attributed to a large volume of deadlifts (kettlebell ladder). The participant continued in the program in a reduced capacity before withdrawing in week seven. Two females experienced an intercostal strain, one with onset of symptoms reported during kettlebell swings of an undisclosed weight, and one while performing a 1RM kettlebell deadlift. Both participants were able to continue training and avoid aggravating exercises, with symptoms resolving consistent with natural history. One female experienced non-traumatic shoulder pain which limited overhead activity. She was able to continue training, however her symptoms did not resolve before the end of the trial. Imaging revealed no tissue or mechanical explanation for the symptoms. Although the trial was not adequately powered to assess safety as an outcome, the absence of any serious adverse events suggests that kettlebell training is likely to have a risk profile similar to other forms of resistance training.

## 4. Discussion

The BELL trial was the first study to examine the effects of 3-months pragmatic hardstyle kettlebell training on grip strength and measures of healthy aging, in insufficiently active apparently healthy older adults. The program had high engagement with few non-serious musculoskeletal events from moderate to high-intensity training, and high training load volume. We believe that frequent supervised training and personalised programming provided in the initial six weeks were key to the safe and effective implementation of community-based group kettlebell exercise for older adults. Kettlebell training resulted in a large clinically important increase in grip strength, with significant improvements in cardiovascular capacity, lean muscle mass, lower limb strength and endurance, and functional capacity. Consistent with previous observation of no non-response to resistance-type exercise in older adults ^(63)^, all participants in the present study demonstrated a positive adaptive response to one or more outcomes.

### 4.1. Grip strength

Handgrip strength decreases with age and is predictive of disability, morbidity, healthcare costs and mortality ^(21, 27, 64)^. At baseline, five females and five males recorded grip strength at or below the 16 kg and 27 kg clinical cut-off values to test for sarcopenia in females and males respectively ^(15)^. For nine of those ten participants, grip strength increased above the clinical cut-off value following training. The estimate of fixed effects pre- to post-training, exceeded the minimum clinically important difference of 5.0 to 6.5 kg ^(62)^. This finding is consistent with findings that the greatest increases in grip strength are observed in higher intensity exercise programs, which involve gripping activities and a high percentage of 1RM ^(65)^. Results from the present study showed improvements in grip strength ≈2x larger than those from a comparable 12-week kettlebell study in older women with sarcopenia (66.7 ± 5 yrs, ASM 15.4 kg, Sarcopenia index 5.57 kg.m^2^) ^(12)^.

### 4.2. Cardiovascular endurance

#### 4.2.1. HR & blood pressure

Age is the primary risk factor for cardiovascular disease, with hypertension a significant modifiable risk factor. Individuals with poor fitness and high resting heart rate (≥80 bpm) have the highest risk of cardiovascular disease and all-cause mortality ^(66)^. Exercise and physical activity can help mitigate the adverse effects ^(67)^ with higher intensity exercise likely to be more beneficial to arterial stiffness than moderate intensity exercise ^(68)^. The 7.4 bpm reduction in resting HR was significant from baseline to 29 weeks, but differences were non-significant between week 13 pre-training and week 29 post-training. Due to changes made to the testing procedures resulting from COVID-19 restrictions, resting blood pressure at week 29 was taken immediately following the DXA scan, with participants lying supine and rested at the start of the test schedule. At all other time points, resting HR was taken at the end of the test schedule in a seated position. Change in resting HR therefore, cannot confidently be attributed to a training effect.

Random effects modelling of dynamic resistance training, show a mean decrease in systolic blood pressure of 1.8 mm Hg 95% CI [3.7 to 0.011] ^(69)^, and in some populations the effects may be comparable to or greater than those achieved with aerobic exercise ^(70)^. The present study however found no significant change in SBP from baseline, although participants were normotensive.

#### 4.2.2. 6-minute walk distance

Designated the “6^th^ vital sign”, walking is used to assess functional status and overall health ^(71)^, with reduced lower limb strength in women associated with slower walking speeds ^(72)^. Conversely, strengthening the lower limbs in older adults may increase walking speed and reduce risk of falls ^(73)^. Faster walking speed and greater walking distance are also associated with significant reductions in cardiovascular disease and all-cause mortality compared to slower, shorter walks, with a more pronounced dose-response effect in those over 50 years of age ^(74, 75)^. At baseline, mean 6MWD for the participants in the present study was 599.8 m, farther than the 486.1 (87.2) m reported by Martien ^(34)^ from 770 community dwelling older adults of a similar age, but within normal age and sex-based reference standards ^(76)^. Shnayderman ^(77)^ reported an improvement in 6MWD of 43.0 m 95% CI [19.6, 68.0] in 26 younger adults (43.6 ± 13.5 years) engaged in 6-weeks of muscle strengthening, a result virtually identical to the 41.7 m (8.7) in the present study. A 7.0% improvement in 6MWD pre-to post-training is suggestive of a training effect, however, only improvements >50 m are likely to exceed a minimum clinically important difference ^(78)^.

#### 4.2.3. Stair climb

Stair climbing is an essential functional activity for independent community-dwelling older adults, with gait speed strongly predictive of performance ^(79)^. Stair climb performance can be improved with lower limb resistance training and stretching exercise ^(80)^, with stair climb time used to estimate aerobic capacity ^(32)^. In a review, Ozaki ^(81)^ reported six of nine studies showing significant improvement in VO2 max in older adults engaged in resistance training, however, such improvements in VO2 may only be likely among individuals with low baseline fitness i.e. a VO2 max under 25ml.kg.min^-1^. Kalapotharakos ^(82)^ reported improvements in VO_2_max from 6.6% to 30%, with effects dependent upon a wide range of factors including individual characteristics and program variables. In the present study, there was a large difference at baseline in the mean VO2 estimated from the stair climb (39.8 ml.kg^-1^.min^-1)^) and the age-predicted maximum (23.8 ml.kg^-1^.min^-1^). The age-predicted maximum is within the reference range for health older adults ^(83)^, thus VO2 estimated from the stair climb, which was considerably higher, was deemed unreliable. Linear mixed effects modelling demonstrates a change in estimated VO_2_max of 4.9ml.kg^-1^.min^-1^, or 12.3%. A 12.3% increase applied to the age-predicted VO_2_max at baseline, suggests a more reliable training effect improvement of 2.9 ml.kg^-1^.min^-1^. Although beneficial, a change less than 5ml.kg^-1^.min^-1^ is likely less than the minimum clinically important difference for healthy older adults ^(84)^.

### 4.3. Muscular strength, power, and endurance

#### 4.3.1. Knee extension strength

Knee extension force is a useful measure to evaluate lower limb function in older adults, particularly in relation to functional activities such as rising from a chair and ascending stairs ^(85)^. Assessment of knee extension strength is recommended as part of a comprehensive geriatric assessment, with physical, nutritional and psychological health characteristics more strongly associated with knee extension strength than handgrip strength ^(86)^ and a better predictor of functional performance than handgrip in older adults in assisted living facilities ^(34)^. At baseline, knee extension force normalised to bodyweight was 40.0 (10.6) % and 43.6 (12.4) % for females and males respectively. These are comparable to population reference data ^(34, 87)^ and higher than functionally relevant cut-off values of 31% and 40% for females and males respectively ^(34)^. Consistent with other studies ^(88, 89)^, leg extension force in the present study increased 19.1% and 17.4% in the right and left legs, respectively pre- to post-training.

#### 4.3.2. Knee extension RFD and lower body power

For older adults, RFD is associated with reduced postural balance and impaired balance recovery after tripping. Age-associated reduction in RFD can be attenuated by life-long resistance training ^(90)^, although responses to training are highly individual ^(91)^. Lower limb RFD may also be a better predictor of mobility in older adults than hand grip strength and sit-to-stand performance ^(92)^. In healthy older adults, improvements in RFD have been reported from explosive and heavy resistance training ^(88)^ and low-repetition power training with a weighted vest ^(93)^. In order to improve RFD, the speed of movement may not be as important as the *intent* to move rapidly ^(94)^. A distinguishing feature of the hardstyle kettlebell swing, is the *intent* to perform the exercise rapidly, that is, the ability to develop force quickly during a rapid voluntary contraction from a low or resting level, thus there is merit to the claim that the ballistic hardstyle swing may improve lower limb RFD ^(4)^. Furthermore, a vertical jumping motion is recommended as a prerequisite exercise for learning the movement pattern of a hardstyle swing. One might expect therefore, that training the swing would improve vertical jump performance, having trained the movement with the intent to do so rapidly. If the present study had been powered to cover all variables, these results would not support that hypothesis, with pre- and post-training comparison showing non-significant changes in lower limb power measured by sit-to-stand performance or vertical jump height.

#### 4.3.3. Hip extension strength and RFD

Reduced hip extension strength may compromise postural balance and increase falls risk in older adults ^(95)^. Mean peak torque of the hip extensors at baseline in the present study, was higher than previously reported ^(36, 95, 96)^ however, peak force was lower than more recent data reporting 227.2 (56.7) N ^(97)^. Pre- to post-training comparison showed a 6.7% increase in hip extension peak force in the right hip and 12.7% in the left hip, which is small but encouraging.

#### 4.3.4. 30sSTS

Rising from a seated position is an essential functional movement for older adults which can be impacted by age and age-associated conditions. Performing too few sit-to-stand manoeuvrers throughout a day can contribute to strength impairment and limitations in physical activity, with 45 recommended as a minimum ^(98)^. Mean 30sSTS in the present study was 14.6 rises, ‘average’ for males and females 70-79 years of age ^(99)^, and similar to the 13.97 reported in a similar cohort ^(100)^. The mean difference pre- to post-training was an improvement of 3.3 reps, or 23%, exceeding the minimum clinically important difference of 2.0-2.6 repetitions for patients with hip osteoarthritis ^(101)^.

### 4.4. Flexibility

#### 4.4.1. Fingertip to floor

Multi-component exercises intended to improve flexibility, are recommended for older adults, however, the most effective intervention characteristics of exercise type, frequency, duration and intensity are unclear ^(102)^. The efficacy and utility of flexibility as a major component in exercise prescription for most populations has been recently questioned ^(103)^, with conflicting information in older adults regarding the relationship between flexibility (and interventions to improve it), and performance in functional activities. A recent systematic review and meta-analysis ^(104)^ suggests that static stretching is not necessary to improve flexibility, and resistance training programs might provide similar outcomes. Although not statistically significant, pairwise comparison pre- to post-training in the present study showed a mean difference of 4.1 cm in the fingertip to floor measure, suggestive of an improvement, although this should be interpreted with caution ^(22)^.

### 4.5. Body composition (DXA)

At baseline, five females had a height-adjusted SMI below 6.0 kg/m^2^, with two individuals below the EWGSOP2 sarcopenia cut-off point of 5.5 kg/m^2 (15)^. Four males at baseline had a SMI below 7.5 kg/m^2^, with one below the 7.0 kg/m^2^ cut-off. Muscle mass decreases with age, but age explains less than 25% of the variance in strength ^(105)^. Losses in lower limb lean mass specifically are related to compromised functional activities. Mean appendicular skeletal muscle mass at baseline of participants in the present study (17.17 kg and 25.23 kg for females and males respectively) lay within the reference ranges for age- and sex-matched Australians ^(106)^. Mean skeletal muscle index (ASM/m^2^) values although marginally lower, were within the expected range; differences might be explained by stature, with shorter individuals more likely to have a lower SMI if not adjusted for height ^(107)^. Magnitude of change in ASM pre- to post-training in the present study (687.9 g), was >2.5x the reported effect size from 8 weeks of moderate to hard intensity kettlebell training in sarcopenic women 65-75 yrs (MD = 260g, δ = 0.11) ^(12)^. This might be explained by the sarcopenic status of participants in the study by Chen, or perhaps difference in training variables. The findings of the present study are contrary to reports of limited low-quality evidence that resistance training is an effective intervention for improving muscle mass in older adults with sarcopenia ^(108)^. In community-dwelling older adults, fat mass is independently associated with a greater decline in HRQoL ^(109)^. Data from the present study suggests that kettlebell training in isolation would not be an effective intervention for reducing adiposity among older adults however, consistent with previous data ^(110)^, results from the present study suggest that frequent moderate to high intensity kettlebell training could be effective for increasing lean mass. While BIA-derived ASM has been reported in the present study, BIA data was deemed to be unreliable.

### 4.6. Functional capacity

#### 4.6.1. 5-times floor transfer

Ability to rise from the floor is a clinical measure of musculoskeletal fitness proposed to predict all-cause mortality ^(111)^, with those unable to rise having a markedly higher risk of having a fall-related injury ^(112)^. Evaluation of floor transfer capacity is a reliable measure of older adults’ functional mobility and recommended as a standardised component of a geriatric physical assessment ^(52, 53)^. The large 14.3% pre-to post-training reduction in 5x floor transfer time was very encouraging. Due to challenges teaching the Turkish get-up (a structured floor transfer manoeuvre specific to kettlebell training) effectively to a class of 14-16 older adults, far less time was spent practicing the Turkish get-up exercise than had initially been planned. As no other exercises were expected to provide specific transference to the floor transfer test, it is proposed that far larger improvements in floor transfer might be expected with greater emphasis given to teaching the Turkish get-up to older adults.

#### 4.6.2. Predicted 1RM deadlift

High intensity (>80% 1RM) resistance training programs are recommended to counter sarcopenia and osteopenia in older adults ^(113, 114)^. Machine-based 1RM measures have been shown to be safe and highly reproducible for older adults, with minimal detectable changes of 1-3% (<1kg) ^(115)^, but prediction equations consistently underestimate 1RM ^(116)^. Knowing or calculating 1RM, may have very limited utility for most people ^(117)^ but its use in clinical practice and research is likely to continue ^(118-120)^. In the present study, participants’ maximal or predicted maximal 1RM kettlebell deadlift did not influence training loads used during the intervention. Change in predicted 1RM was not used as a proxy for change in strength, rather it was chosen as an evaluation of the participant’s functional capacity to perform a maximal lift ^(121)^. This is of particular interest for healthcare providers working with older adults who may have developed maladaptive cognitions relating to their actual or perceived capacity to safely lift a heavy object. ^(122, 123)^. Performing a large training volume with kettlebells up to 80kg, it was expected that their physical capacity would significantly change, so the moderate pre- to post-training increase of 16.2 kg (23.3%) was unsurprising. Greater improvements may have been anticipated if the participants had been able to train with the heavier (44-80 kg) kettlebells for the second half of the trial.

### 4.7. Quality of health

#### 4.7.1. SF36 & Sense of Coherence

Handgrip and walking speed are two of the most powerful biomarkers of HRQoL in older adults ^(124)^, highlighting the importance of maintaining physical capacity as a key element in successful aging. Exercise, regardless of type, is associated with lower mental health burden, with aerobic and gym activities, durations of 45 mins, and training frequencies of three to five times per week associated with the largest reductions ^(125, 126)^. Training just twice a week however is likely to improve QoL and Sense of Coherence ^(127)^. Physical activity improves happiness and mood (positive affect), self-efficacy and self-esteem in the long-term ^(128)^, although this may differ for frail older adults ^(129)^. Self-efficacy is a significant mediator for older adults engaging in healthy lifestyle activities ^(130)^, with resistance training specifically having been shown to significantly improve HRQoL with greatest effect in the domains of mental health and bodily pain ^(131)^. In the present study, overall *health change* was the only sub-domain of the SF-36 to improve significantly, with no significant change in Sense of Coherence. This may have reflected the disruption and anxiety caused by the concurrent arrival of SARS-CoV2 in Australia, and COVID-19 restrictions which prevented the continuation of face-to-face training mid-way through training.

The salutogenic concept of Sense of Coherence ^(58)^, which is the individual’s perceived control over and ability to improve their physical, mental, and social health and wellbeing, has been shown to predict HRQoL ^(132)^. Health promotion strategies (should) create environments which empower people to identify and make use of their own resources to this end. Providing older adults with the knowledge, practical skills, and group training opportunities to use kettlebells appears to be a safe and effective health promotion strategy.

### 4.8. Training load

Hardstyle kettlebell training does not follow all of the traditional resistance training protocols in relation to sets, repetitions, loads, or rest periods. It does however involve a significant within-session training load volume, which is beneficial for strength and hypertrophy ^(133)^. High volume training may be advantageous or necessary in some cases ^(134, 135)^, but improvements were still anticipated among participants with the lowest training load volume, as low volume resistance training can still improve muscle strength and functional performance in older adults, with no evidence of non-responsiveness ^(136)^. An unexpected finding of the present study was the linear increase in training load after face-to-face training stopped, when participants were required to train at home with limited access to kettlebells. Training load volume was only planned at an individual level during the final two weeks of the training.

## 5. Strengths and limitations

There are several strengths to this study including the unique focus on insufficiently active older adults, high engagement with training, and the large numbers of clinically meaningful outcome measures. This is the first study to assess the feasibility of engaging older adults in a pragmatic group-based hardstyle kettlebell training to promote healthy aging in the community. The pragmatic design with comprehensive exercise reporting, replicated a training approach which had been used successfully in a Physiotherapy practice with older adults for over 18 months, increasing our confidence that it could be readily replicated with minimal barriers to entry. Functional capacity, experience of and tolerance to structured exercise, was varied among participants at baseline, with several having comorbid health conditions which typically limit involvement in high intensity training. Thus, the participants were representative of the sample population for whom this intervention could be applied. In addition, participant-determined exposure enabled a more appropriate comparison of dose based on self-perceived ratings of perceived exertion, also typical of normal practice. The cost-effective and time-efficient nature of the training provides an attractive alternative to healthcare providers to promote group-based resistance training for older adults. However, several limitations of the present study must be acknowledged. Firstly, the single cohort, repeated measures design has challenges with interval validity, particularly with respect to the absence of blinding and lack of a separate control group. Secondly, the large number of secondary outcome measures raises the likelihood of some changes being a false positive observation, and results from secondary outcomes for which the study was not powered, should be interpreted with caution. Finally, the PRESCIS-2 domains of ‘recruitment’ and ‘setting’ scored low (supplementary data) and may influence external validity. Program delivery from a community or clinic setting, operating within an environment or framework governed or influenced by a myriad of factors, such as clinical traditions, health service organisation, staffing and resources constraints, and funding arrangements, may influence quantitative and qualitative outcomes. Ultimately, the effect and success of a program may be significantly influenced by the local framework of healthcare driver domains, including the choice to participate and the Instructor-participant interaction ^(137)^. Linear mixed effect modelling however, increases our confidence that the results can be generalised to a random sample of participants with similar characteristics.

## 6. Conclusion

In conclusion, our findings demonstrate that group-based hardstyle kettlebell training performed at moderate to high-intensity, can be used safely and effectively to engage insufficiently active older adults in the community and promote healthy aging. Insufficiently active males and females 60-80 years of age were able to train 5 days weekly for 3-months, and maintain a very high level of engagement, with no serious adverse events and very low dropout rate. Participants developed the confidence and skills to train independently at home, with large improvements in grip strength, and significant changes in cardiovascular capacity, muscular strength and endurance, functional capacity, and body composition. Further investigations are warranted to determine optimal exercise prescription for insufficiently active older adults with different functional needs and varying physical capacity.

## Supporting information

CERT checklist

CONSORT checklist

PRECIS-2 summary

SPIRIT checklist

TIDieR checklist

Training program

Schedule of data collection

## Data Availability

Data will be made available in accordance with Bond University Research Data Management and Sharing Policy (TLR 5.12, Sections 9.3, 10.2. 10.5, 10.8). Research data and primary materials will be made openly available for use by other researchers and interested persons for further research after a reasonable period following completion of the research.

## CRediT authorship contribution statement

**Neil Meigh:** Conceptualisation, investigation, methodology, project administration, resources, supervision, visualisation, writing - original draft, writing - review & editing, **Justin Keogh**: Conceptualisation, methodology, supervision, writing - review & editing, **Ben Schram**: Conceptualisation, writing - review & editing, **Wayne Hing**: Conceptualisation, methodology, supervision, writing - review & editing, **Evelyne Rathbone**: provided statistical expertise in clinical trial design, data analysis and interpretation. All authors read and approved the final manuscript.

## Acknowledgements

The authors would like to sincerely thank the participants who took part in the trial for their valuable contribution, and the volunteers who assisted with data collection for without whom, this study could not have been completed.

## Registration

Australian New Zealand Clinical Trials Registry (ACTRN12619001177145).

## Protocol

The study protocol was published to the Open Science Framework: https://osf.io/dz96p/.

## Supplementary data

The following supplementary data are available:

- Schedule of data collection
- SPIRIT Checklist
- TIDieR Checklist
- PRECIS-2 Statement
- CERT Checklist
- CONSORT Checklist
- Training program

Supplementary data are available under the terms of the <u>Creative Commons Zero “No rights reserved” data waiver</u> (CC0 1.0 Public domain dedication).

## Abbreviations

1RM: 1 Repetition Maximum
30sSTS: 30-Second Sit-to-Stand
6MWD: 6-Minute Walk Distance
6MWT: 6-Minute Walk Test
AU: Arbitrary Unit
BIA: Bioelectrical Impedance Analysis
BP: Blood Pressure
bpm: beats per minute
CMCJ: Counter-movement vertical jump
COP: Centre of Pressure
DOMS: Delayed-Onset Muscle Soreness
DXA: Dual-energy X-ray absorptiometry
FTT: Floor Transfer Test
5xFT: 5-times Floor Transfer
FTF: Fingertip-to-Floor
HRPF: Health-related Physical Fitness
HRQoL: Health-Related Quality of Life
QoL: Quality of Life
RFD: Rate of Force Development
RKC: Russian Kettlebell Certification
RPE: Rating of Perceived Exertion
RT: Resistance Training
SCT: Stair-climbing Test
SD: Standard Deviation
SoC: Sense of Coherence
sRPE: Session Rating of Perceived Exertion
STS: Sit to Stand
TL: Training Load
V-TL: Training Load Volume

