## Supplementary material for "Effects of supervised high-intensity hardstyle kettlebell training on grip strength and health-related physical fitness in insufficiently active older adults: The BELL pragmatic controlled trial": CERT checklist

**The BELL (Ballistic Exercise of the Lower Limb) trial: A repeated measures, single cohort, pragmatic hardstyle kettlebell training program to improve grip strength, health-related physical fitness, and quality of life in sedentary older adults.**

Australian New Zealand Clinical Trials Registry (ID: ACTRN12619001177145).

**CERT (Consensus on Exercise Reporting Template) Checklist**

| Section/Topic | Item | Checklist item | Addressed on page number/ heading |
|---|---|---|---|
| WHAT: materials | 1 | Detailed description of the type of exercise equipment (e.g., weights, exercise equipment such as machines, treadmill, bicycle ergometer etc)<br>• 4 - 80 kg kettlebells. 8 kg kettlebell used for home exercise Week 1-6. Available weights (8-40kg) distributed to participants during COVID-19 shut-down; each participant provided with two additional kettlebells (not everyone received the same weights). | 2.4. |
| WHO: provider | 2 | Detailed description of the qualifications, teaching/supervising expertise, and/or training undertaken by the exercise instructor.<br>• Exercise instruction will be provided by a single hardstyle-certified (RKC) kettlebell instructor; a Physiotherapist (D.Phty), *Exercise Scientist (BScEx) and Personal Trainer (CertIV Fit) with >25 years resistance training experience, 7 years hardstyle kettlebell training and >18-months running group kettlebell classes using the same training principles and practices*.* | 2.4. |
| HOW: delivery | 3 | Describe whether exercises are performed individually or in a group.<br>• Participants attended the Bond University High Performance Training Centre, Gold Coast, Australia, three-times weekly (Mon, Wed, Fri), for 45-min group classes (*n* ≤16), and prescribed home exercises performed individually twice-weekly (Mon, Thur). | 2.4. |
| | 4 | Describe whether exercises are supervised or unsupervised and how they are delivered.<br>• Group exercise was delivered and supervised face-to-face (weeks 1-6). All exercise was performed at home from week 7 onwards due to the COVID-19 pandemic and local restrictions. Home exercise was unsupervised. *Participants were provided with video links and received frequent (typically daily) written and video updates via private Facebook group and/or email.** | 2.4. |
| | 5 | Detailed description of how adherence to exercise is measured and reported.<br>• A daily training record was captured for analysis. Paper records were used during weeks 1-6, collected by the lead examiner at the end of each session and transcribed to a database for analysis. During weeks 7-12, daily training records (Mon-Fri) were submitted via Survey Monkey for analysis. 'Group session' training data included training load volume (exercises, sets, reps), compliance with prescribed home exercise was Y/N (completed or not completed). | 2.4.<br>2.6.10 |

| | | | |
|---|---|---|---|
| | 6 | Detailed description of motivation strategies<br>• *The Instructor took part in (some of) the training in leading and demonstrating*. * Participants received frequent individual and group encouragement, both publicly and privately. Recognition was given to overcoming challenges, extraordinary effort, and achieving a 'personal best'. Training and communication promoted group engagement to foster a spirit of support, camaraderie, and healthy competition. A private Facebook page was heavily used to provide encouragement, maintain accountability, and foster a community spirit. Participants were encouraged to make use of the on-site coffee-shop after training. | 2.6.10 |
| | 7a | Detailed description of the decision rule(s) for determining exercise progression.<br>• During the first two weeks, participants were advised to work at a relatively low intensity (2-4/10: "easy" to "somewhat hard") with a low volume training load. From Week 3 onward, participants were encouraged to work up to a session-Rate of Perceived Exertion (sRPE) of 5-7/10 (described as "hard" to "very hard") as tolerated. Maximal effort (9-10/10) was discouraged. Where technique was acceptable and RPE appeared to be <4/10, participants were encouraged to increase exercise intensity (kettlebell weight). Exercises were modified where necessary to account for physical limitations or emergent biopsychosocial factors. Exercises were adjusted as necessary to accommodate the physical limitations, stage of learning, and progression of each participant, and participants were able to self-select weights and change any program variable within the group sessions. *Exercise progression (in volume or intensity) was a pragmatic decision based upon the Instructor's sense of safety, competency, physical capacity and desire of the participant or group to complete a given task, in conjunction with the participant's willingness to progress. ** | 2.4. |
| | 7b | Detailed description of how the exercise program was progressed.<br>• The program goal was to steadily increase training volume and intensity as tolerated, while maintaining a participant-reported sRPE which remained moderate to high. Training sessions were planned based on a) physical capacity of the group, b) participant feedback, c) intent to offer variety, and d) plan to progress skill, intensity, and training load volume throughout the trial, and were prepared within the preceding 36hrs. Training load volume was only planned at an individual level during the final two weeks of the training so that (all) participants were able to attained a personal best session V-TL on the final (official) day of the trial. | 2.4.<br>4.8 |
| | 8 | Detailed description of each exercise to enable replication (e.g., photographs, illustrations, video etc.) | Supplementary data |
| | 9 | Detailed description of any home program component (e.g., other exercises, stretching etc.) | Supplementary data |
| | 10 | Describe whether there are any non-exercise components (e.g., education, cognitive behavioural therapy, massage etc) | N/A |
| | 11 | Describe the type and number of adverse events that occurred during exercise.<br>• 4 non-serious adverse events. | 3.3.8. Adverse events |
| WHERE: location | 12 | Describe the setting in which the exercises are performed.<br>• Group exercise took place in the High Performance Training Centre, at Bond University Institute of Health and Sport, Robina, Queensland (AUS). | 2.4. |
| WHEN, HOW MUCH: dosage | 13 | Detailed description of the exercise intervention including, but not limited to, number of exercise repetitions/sets/sessions, session duration, intervention/program duration etc. | Supplementary data |

| | | | |
|---|---|---|---|
| TAILORING: what, how | 14a | Describe whether the exercises are generic (one size fits all) or tailored whether tailored to the individual<br>• Group training was one size fits all insofar as all participants were asked to perform the same exercises however, exercises were modified where necessary to account for physical limitations or emergent biopsychosocial factors e.g., anxiousness lifting overhead. Furthermore, participants were able to self-select weights (intensity) and change any program variable or not perform an exercise if they were not comfortable. | 2.4. |
| | 14b | Detailed description of how exercises are tailored to the individual<br>• Inclusion criteria required that participants had the requisite physical capacity to be able to participate fully. *Most kettlebell exercises can be tailored or made easier by using a lighter weight. Participants with painful arthritis knees were able to manage symptoms by limiting range of knee flexion. ** | 2.3. |
| | 15 | Describe the decision rule for determining the starting level at which people commence an exercise program (such as beginner, intermediate, advanced etc.)<br>• As a skilled mode of dynamic resistance exercise, all participants were considered beginners. *Training was delivered with the following framework in mind: 1) Skill acquisition (with or without load), 2. Practice (characterized by a higher volume at lower intensity), and 3) Train (characterised by a lower volume at higher intensity). ** | N/A |
| HOW WELL: planned, actual | 16a | Describe how adherence or fidelity to the exercise intervention is assessed/measured.<br>• Exercise adherence was reported and recorded daily, with 100% attendance defined as completion of 36, 'group' sessions over the 3-month trial period. Compliance with prescribed home exercise was self-reported as complete or not completed (Y/N). 100% attendance and compliance was defined as 60 training sessions completed (group and individual) over the 3-month trial period. | 2.6.11. Attendance, compliance, and adverse events |
| | 16b | Describe the extent to which the intervention was delivered as planned.<br>• Exercises and V-TL were not set a-priori as participant's physical capacity was unknown to investigators. *Two exercises which could not be taught as anticipated were the snatch and Turkish get-up. The Turkish get-up (a structured floor transfer) was found to be too challenging to teach effectively within the large group setting, resulting in it being largely left to be practiced at home. Accumulation of training load volume was given a higher priority than the TGU, with far less time spent practicing is than had been hoped for. The snatch was used less than anticipated due to similar challenges with teaching and learning it effectively in a large group with heterogenous physical competency. ** | 2.6.10. |

* *Details not provided in the manuscript.*
