## Supplementary material for "Effects of supervised high-intensity hardstyle kettlebell training on grip strength and health-related physical fitness in insufficiently active older adults: The BELL pragmatic controlled trial": CONSORT checklist

**The BELL (Ballistic Exercise of the Lower Limb) trial: A repeated measures, single cohort, pragmatic hardstyle kettlebell training program to improve grip strength, health-related physical fitness, and quality of life in sedentary older adults.**

Australian New Zealand Clinical Trials Registry (ID: ACTRN12619001177145).

**CONSORT Checklist of items for reporting pragmatic trials**

| Section | Item | Standard CONSORT description | Extension for pragmatic trials | Addressed on page number/ heading |
|---|---|---|---|---|
| Title and abstract | 1 | How participants were allocated to interventions (e.g., “random allocation,” “randomised,” or “randomly assigned”) | | N/A |
| **Introduction** | | | | |
| Background | 2 | Scientific background and explanation of rationale | Describe the health or health service problem that the intervention is intended to address and other interventions that may commonly be aimed at this problem | 2, 1. Introduction |
| **Methods** | | | | |
| Participants | 3 | Eligibility criteria for participants; settings and locations where the data were collected | Eligibility criteria should be explicitly framed to show the degree to which they include typical participants and/or, where applicable, typical providers (e.g., nurses), institutions (e.g., hospitals), communities (or localities e.g., towns) and settings of care (e.g., different healthcare financing systems) | 2.3. Participants 5. Strengths and limitations |
| Interventions | 4 | Precise details of the interventions intended for each group and how and when they were actually administered | Describe extra resources added to (or resources removed from) usual settings in order to implement intervention. Indicate if efforts were made to standardise the intervention or if the intervention and its delivery were allowed to vary between participants, practitioners, or study sites<br><br>Describe the comparator in similar detail to the intervention | 2.4. Exercise intervention, 2.5. Control activities Supplementary data |
| Objectives | 5 | Specific objectives and hypotheses | | 1. Introduction |
| Outcomes | 6 | Clearly defined primary and secondary outcome measures and, when applicable, any methods used to enhance the quality of measurements (e.g., multiple observations, training of assessors) | Explain why the chosen outcomes and, when relevant, the length of follow-up are considered important to those who will use the results of the trial | 2.1. Study design and sample size |
| Sample size | 7 | How sample size was determined; explanation of any interim analyses and stopping rules when applicable | If calculated using the smallest difference considered important by the target decision maker audience (the | 2.1. Study design and sample size |

| Section | Item | Standard CONSORT description | Extension for pragmatic trials | Addressed on page number/ heading |
|---|---|---|---|---|
| | | | minimally important difference) then report where this difference was obtained | |
| Randomisation—sequence generation | 8 | Method used to generate the random allocation sequence, including details of any restriction (e.g., blocking, stratification) | | N/A |
| Randomisation—allocation concealment | 9 | Method used to implement the random allocation sequence (e.g., numbered containers or central telephone), clarifying whether the sequence was concealed until interventions were assigned | | N/A |
| Randomisation—implementation | 10 | Who generated the allocation sequence, who enrolled participants, and who assigned participants to their groups | | N/A |
| Blinding (masking) | 11 | Whether participants, those administering the interventions, and those assessing the outcomes were blinded to group assignment | If blinding was not done, or was not possible, explain why | 2.1. Study design and sample size |
| Statistical methods | 12 | Statistical methods used to compare groups for primary outcomes; methods for additional analyses, such as subgroup analyses and adjusted analyses | | 2.7. Statistical analysis |
| **Results** | | | | |
| Participant flow | 13 | Flow of participants through each stage (a diagram is strongly recommended)—specifically, for each group, report the numbers of participants randomly assigned, receiving intended treatment, completing the study protocol, and analysed for the primary outcome; describe deviations from planned study protocol, together with reasons | The number of participants or units approached to take part in the trial, the number which were eligible, and reasons for non-participation should be reported | Fig. 4 Participant flow |
| Recruitment | 14 | Dates defining the periods of recruitment and follow-up | | Fig .1 Study timeline |
| Baseline data | 15 | Baseline demographic and clinical characteristics of each group | | 3.1 Participant characteristics at baseline |
| Numbers analysed | 16 | Number of participants (denominator) in each group included in each analysis and whether analysis was by “intention-to-treat”; state the results in absolute numbers when feasible (eg, 10/20, not 50%) | | 2.7. Statistical analysis<br>3. Results |
| Outcomes and estimation | 17 | For each primary and secondary outcome, a summary of results for each group and the estimated effect size and its precision (e.g., 95% CI) | | Table 3<br>Table 4 |

| Section | Item | Standard CONSORT description | Extension for pragmatic trials | Addressed on page number/ heading |
|---|---|---|---|---|
| Ancillary analyses | 18 | Address multiplicity by reporting any other analyses performed, including subgroup analyses and adjusted analyses, indicating which are prespecified and which are exploratory | | N/A |
| Adverse events | 19 | All important adverse events or side effects in each intervention group | | 3.3.8. Adverse events |
| **Discussion** | | | | |
| Interpretation | 20 | Interpretation of the results, taking into account study hypotheses, sources of potential bias or imprecision, and the dangers associated with multiplicity of analyses and outcomes | | 4. Discussion<br>5. Strengths and limitations |
| Generalisability | 21 | Generalisability (external validity) of the trial findings | Describe key aspects of the setting which determined the trial results. Discuss possible differences in other settings where clinical traditions, health service organisation, staffing, or resources may vary from those of the trial | 5. Strengths and limitations |
| Overall evidence | 22 | General interpretation of the results in the context of current evidence | | 4. Discussion |

***Cite as:** Zwarenstein M, Treweek S, Gagnier JJ, Altman DG, Tunis S, Haynes B, Oxman AD, Moher D for the CONSORT and Pragmatic Trials in Healthcare (Practihc) group. Improving the reporting of pragmatic trials: an extension of the CONSORT statement. BMJ 2008; 337;a2390.*
