## Supplementary material for "Effects of supervised high-intensity hardstyle kettlebell training on grip strength and health-related physical fitness in insufficiently active older adults: The BELL pragmatic controlled trial": PRECIS-2 summary

**The BELL (Ballistic Exercise of the Lower Limb) trial: A repeated measures, single cohort, pragmatic hardstyle kettlebell training program to improve grip strength, health-related physical fitness, and quality of life in sedentary older adults.**

Australian New Zealand Clinical Trials Registry (ID: ACTRN12619001177145).

**The PRagmatic-Explanatory Continuum Indicator Summary 2 (PRECIS-2)**

PRECIS-2 scores for trial domains

| | Domain | Score | Rationale |
|---|---|---|---|
| 1 | Eligibility criteria | 5 | Exclusion criteria precluded only those who would not be suitable candidates to participate in usual practice - a representative cohort of the (insufficiently active) local community, and ‘usual care’ for community-based group-exercise programs; people are free to participant if physically able and safe to do so. |
| 2 | Recruitment Path | 2 | An explanatory approach with targeted invitations via print media; not an existing kettlebell training program, community group-exercise facility or clinical practice however, print media may be used to promote community-based group-exercise programs for older adults. |
| 3 | Setting | 3 | Specialised trial to be conducted from a single site (large gymnasium), likely to be shared with a small (n<10) group of athletes. Not a typical setting for existing community group-exercise programs, however gymnasiums would likely be used due to equipment requirements. |
| 4 | Organisation intervention | 4 | An explanatory approach however, organisation, complexity, staff levels and training delivery are representative of usual practice. Trainer requires kettlebell training (more than usual experience or certification). |
| 5 | Flexibility of experimental intervention (delivery) | 5 | Pragmatic choice in delivery of the intervention. Participants provided with guidelines of goal attainment and required to monitor load, however no measures to improve compliance beyond usual practice. Trial closely replicated a previously delivered clinic-based program. |
| 6 | Flexibility of experimental intervention (adherence) | 5 | No more than usual encouragement to adhere to the intervention. No exclusion based on adherence or measures to improve adherence. Participants required to complete training diary. No measures to improve compliance beyond typical strategies for group-exercise programs. |
| 7 | Follow up | 5 | Participants will not be followed-up beyond immediate data collection at completion of the 12-week training program. No more than usual practice. |
| 8 | Outcome | 5 | Primary outcome(s) are directly relevant to healthcare providers. Relationship to physical and functional capacity is obvious to participants. |
| 9 | Analysis | 5 | Linear mixed effects modelling increases our confidence that the results can be generalised to a random sample of participants with similar characteristics. |

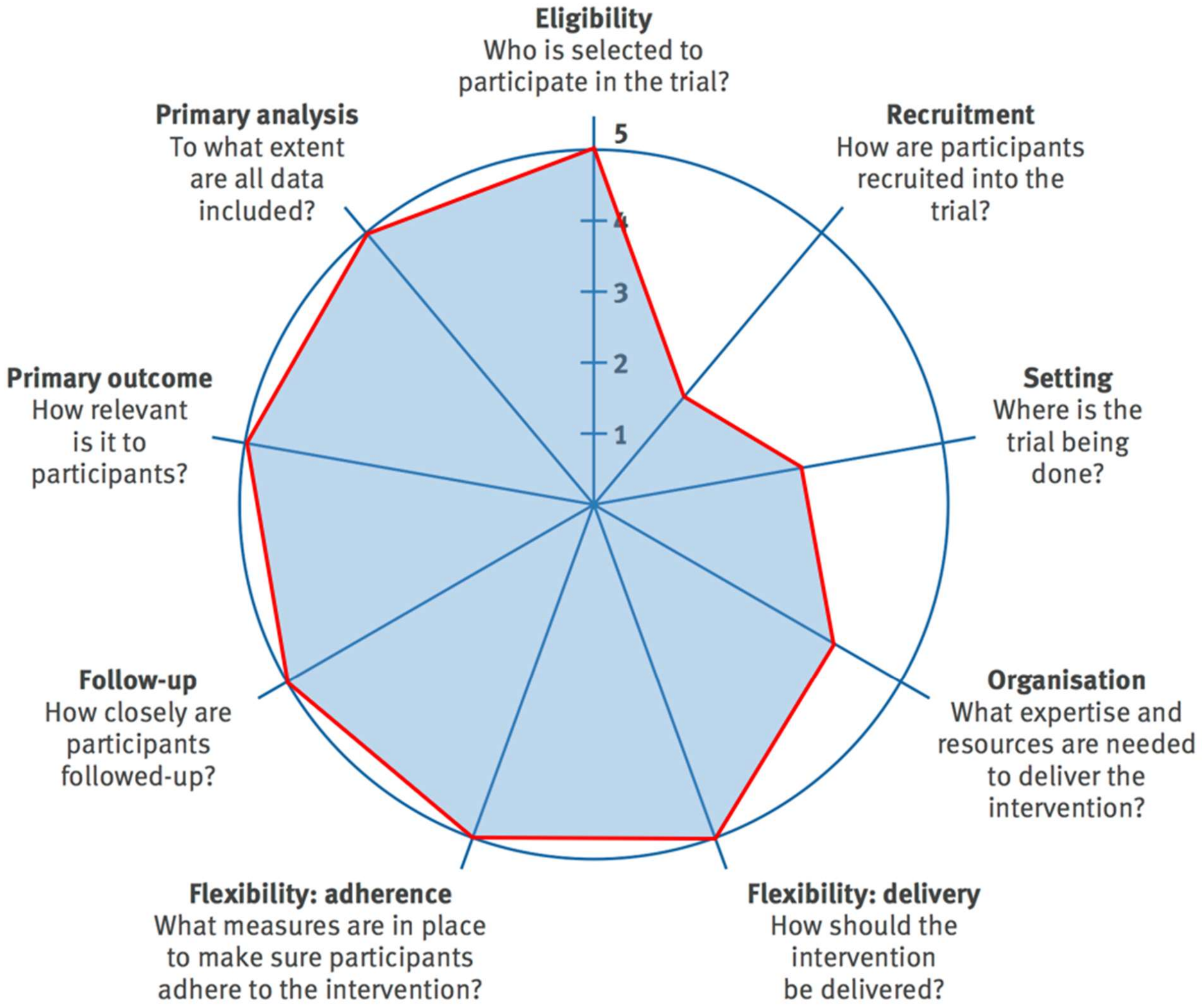


Figure: The PRagmatic-Explanatory Continuum Indicator Summary 2 (PRECIS-2) wheel
