## Supplementary material for "Effects of supervised high-intensity hardstyle kettlebell training on grip strength and health-related physical fitness in insufficiently active older adults: The BELL pragmatic controlled trial": TIDieR checklist

TIDieR
Template for Intervention Description and Replication

### The BELL (Ballistic Exercise of the Lower Limb) trial: A repeated measures, single cohort, pragmatic hardstyle kettlebell training program to improve grip strength, health-related physical fitness, and quality of life in sedentary older adults.

Australian New Zealand Clinical Trials Registry (ID: ACTRN12619001177145).

**The TIDieR (Template for Intervention Description and Replication) Checklist***

| Item | Description | Addressed on page number/ heading |
|---|---|---|
| **BRIEF NAME** | | |
| **1.** | Provide the name or a phrase that describes the intervention.<br>• Group-based hardstyle kettlebell training | Title |
| **WHY** | | |
| **2.** | Describe any rationale, theory, or goal of the elements essential to the intervention. *<br>• Training **frequency:** 5 days/week (group training 3 days + home-training 2 days) with the *rationale* that exercising more frequently will provide better outcomes. Also, to test the *theory* that inactive older adults can tolerate daily training.<br>• Training **intensity**: moderate to high during supervised group classes to maximise mechanical stimulus for physiological adaptation. The goal was to reach the end of the trial with participants feeling comfortable training at a “hard” to “very hard” intensity, with the *theory* that this would help improve physical self-confidence.<br>• Training **independently**: low volume, low intensity training at home was included with the *rationale* that it would i) encourage independent training, ii) increase time spent on skill acquisition (which would in turn improve class performance/outcomes), iii) improving confidence with the equipment, and iv) encouraging greater incidental activity. The *goal* was to increase chronic training load volume and develop a habit of engaging in intentional daily exercise. A low minimum target was set so that participants felt it was achievable (the *rationale* that easy ‘wins’ are motivating), with no upper limit for those who wanted to do more and train harder. Additionally, it was hoped that training at home would foster a sense safety and not needing to be reliant upon an Instructor.<br>• Training **volume:** The *goal* was to maintain a steady increase in training load volume throughout the trial.<br>• **Hardstyle**: Programming was based upon the fundamental hardstyle techniques of, kettlebell swing, clean, press, squat, snatch, and Turkish get-up. The *goal* was to test whether an approach previously used in practice, predominantly with younger adults (20-50yrs), could be used successfully with an older cohort. The *rationale* for using hardstyle is that it is the most widely recognised “style” of kettlebell training, it offers a clear and scalable working framework for Instructors (and participants) which is based on sound principles, and its techniques can be more objectively measured and compared between sites/providers/individuals.<br>• **Routine practice**: The *goal* was no to treat the participants any differently because they were older. Exercises were modified and scaled as necessary however, the *rationale* was that if the participants were treated as capable and competent by the Instructor (and Physiotherapist), they would be more likely to see themselves that way and engage in the most desirable way. The *theory* was that | 2.4. Exercise intervention * |

participants would be more likely to respond positively if they felt encouraged rather than limited/restricted based upon a perceived notion of age-limited capacity or risk.

- **Dose**: Older adults are routinely underdosed in clinical practice. The *goal* was to challenge common training approaches; mode of exercise, intensity (weight), training volume, and effort (RPE).
- **Technique**: Exercises were progressed based on the Instructor's judgement of safety for a given individual, and the person's desire to progress. Less emphasis than is typical of hardstyle practice was given to "perfecting" technique. Progression in weight or volume was permitted if it appeared safe to do so and the participant felt comfortable. The *rationale* was that ideal technique may not be achievable due to individual physical limitations or insufficient time (and one-on-one instruction) to achieve the desired outcome. The *theory* is that "ideal" technique is not a prerequisite for progression or predictive of better outcomes, and allowing progression with less-than-ideal technique would provide greater psychosocial benefit to the individual within the group setting, as opposed to unnecessarily holding someone back. The *goal* was for participants to have fun and enjoy the training, (which accumulating a significant training load volume), not to become kettlebell experts.
- Instructor **engagement**: The Instructor would be immersed in the delivery. Previous data highlights the value participants in community-based group-exercise programs have placed on the Instructor-participant relationship, and the Instructor's personal characteristics. Social interaction in the training process would be maximised wherever possible, including participating in the training when feasible to do so, with the *rationale* that this would help improve engagement and compliance.

**WHAT**

| | | |
|---|---|---|
| **3.** | Materials: Describe any physical or informational materials used in the intervention, including those provided to participants or used in intervention delivery or in training of intervention providers. Provide information on where the materials can be accessed (e.g. online appendix, URL).<br>• Training program with video links provided as supplementary data | 2.4. Exercise intervention Supplementary data |
| **4.** | Procedures: Describe each of the procedures, activities, and/or processes used in the intervention, including any enabling or support activities.<br>• *The Instructor took part in (some of) the training in leading and demonstrating.* * Participants received frequent individual and group encouragement, both publicly and privately. Recognition was given to overcoming challenges, extraordinary effort, and achieving a 'personal best'. Training and communication promoted group engagement to foster a spirit of support, camaraderie, and healthy competition. A private Facebook page was heavily used to provide encouragement, maintain accountability, and foster a community spirit. Participants were encouraged to make use of the on-site coffee-shop after training. | 2.6.11. Attendance, compliance, and adverse events |

**WHO PROVIDED**

| | | |
|---|---|---|
| **5.** | For each category of intervention provider (e.g. psychologist, nursing assistant), describe their expertise, background and any specific training given.<br>• Exercise instruction will be provided by a single hardstyle-certified (RKC) kettlebell instructor; a Physiotherapist (D.Phty), *Exercise Scientist (BScEx) and Personal Trainer (CertIV Fit) with >25 years resistance training experience, 7 years hardstyle kettlebell training and >18-months running group kettlebell classes using the same training principles and practices*.* | 2.4. Exercise intervention CERT checklist |

**HOW**

| | | |
|---|---|---|
| **6.** | Describe the modes of delivery (e.g. face-to-face or by some other mechanism, such as internet or telephone) of the intervention and whether it was provided individually or in a group.<br>• The participants will receive a thorough instruction in the strengthening exercises conducted one-to-one by a physiotherapist. Thus, the instruction is supervised, but hereafter, the exercises are performed individually without supervision. | 2.4. Exercise intervention Supplementary data |

| | | |
|---|---|---|
| **WHERE** | | |
| **7.** | Describe the type(s) of location(s) where the intervention occurred, including any necessary infrastructure or relevant features.<br>• Participants attended the Bond University High Performance Training Centre, Gold Coast, Australia, three-times weekly (Mon, Wed, Fri), for 45-min group classes (n = ≤16), and twice-weekly (Mon, Thur) prescribed home exercises performed individually. Group exercise was delivered and supervised face-to-face (weeks 1-6). All exercise was performed at home from week 7 onwards due to the COVID-19 pandemic and local restrictions. Home exercise was unsupervised. *Participants were provided with video links and received frequent written and video updates via private Facebook group and/or email.** | 2.4. Exercise intervention Supplementary data |
| **WHEN and HOW MUCH** | | |
| **8.** | Describe the number of times the intervention was delivered and over what period of time, including the number of sessions, their schedule, and their duration, intensity or dose.<br>• Attendance and *compliance* were recorded for 12 weeks, with 100% attendance and *compliance* defined as 60 training sessions (group and individual) over the 3-month trial period. Group training sessions were 45 mins in duration, including a mobility warm-up ≈ 5 mins; *Group 1, 09:30 - 10:15, Group 2 10:30 - 11:15. n = 14-16 (max) per group.* * | 2.4. Exercise intervention Fig. 5 Training load volume |
| **TAILORING** | | |
| **9.** | If the intervention was planned to be personalised, titrated or adapted, then describe what, why, when, and how.<br>• During the first two weeks, participants were advised to work at a relatively low intensity (2-4/10: "easy" to "somewhat hard") with a low volume training load. From Week 3 onward, participants were encouraged to work up to a session-Rate of Perceived Exertion (sRPE) of 5-7/10 (described as "hard" to "very hard") as tolerated. Maximal effort (9-10/10) was discouraged. Where technique was acceptable and RPE appeared to be <4/10, participants were encouraged to increase exercise intensity (kettlebell weight). Exercises were modified where necessary to account for physical limitations or emergent biopsychosocial factors. Exercises were adjusted as necessary to accommodate the physical limitations, stage of learning, and progression of each participant, and participants were able to self-select weights and change any program variable within the group sessions. *Exercise progression (in volume or intensity) was a pragmatic decision based upon the Instructor's sense of safety, competency, physical capacity and desire of the participant or group to complete a given task, in conjunction with the participant's willingness to progress*. *<br>• The program goal was to steadily increase training volume and intensity as tolerated, while maintaining a participant-reported sRPE which remained moderate to high. Training sessions were planned based on a) physical capacity of the group, b) participant feedback, c) intent to offer variety, and d) plan to progress skill, intensity, and training load volume throughout the trial, and were prepared within the preceding 36hrs. Training load volume was only planned at an individual level during the final two weeks of the training so that (all) participants were able to attained a personal best session V-TL on the final (official) day of the trial. | 2.4. Exercise intervention |
| **MODIFICATIONS** | | |
| **10.** | If the intervention was modified during the course of the study, describe the changes (what, why, when, and how). | N/A |
| **HOW WELL** | | |
| **11.** | Planned: If intervention adherence or fidelity was assessed, describe how and by whom, and if any strategies were used to maintain or improve fidelity, describe them.<br>• At the end of each group training session, participants submitted a training record of the exercises performed, weights used, number of sets and repetitions completed, and sRPE. During intervention weeks 1-6, a paper record was collected by the lead examiner at the end of each session and transcribed to a database for analysis. During intervention Weeks 7-12, daily training records (Mon-Fri) | 2.4. Exercise intervention 2.6.10. Training load 2.6.11. |

| | | |
|---|---|---|
| | were submitted via Survey Monkey. Training records and attendance\compliance data were collected, entered into a database and analysed by the lead investigator. | Attendance, compliance, and adverse events |
| **12.** | Actual: If intervention adherence or fidelity was assessed, describe the extent to which the intervention was delivered as planned.<br>• Exercises and V-TL were not set a-priori as participant's physical capacity was unknown to investigators. | 2.6.10. Training load |

* Details not explicit in the trial manuscript
