## Supplementary material for "Effects of supervised high-intensity hardstyle kettlebell training on grip strength and health-related physical fitness in insufficiently active older adults: The BELL pragmatic controlled trial": Training program

TIDieR
Template for Intervention Description and Replication

### The BELL Trial: Program schedule

Warm-up sequence: https://youtu.be/6iLRvW-yQbE

#### Week 1

- **Day 1 -** (10/2/20)
  - Farmer's carry - 2 bells (5 rounds) approx. 150m
  - Deadlift x10, 2H-swing x10, rows x10 (4 rounds)
- **Day 2**
  - Wall-assisted TGU, x10 each side: https://youtu.be/G8FI6mu2IYk
- **Day 3 -** (12/2/20)
  - 2x 30s holds in quadruped
  - 1H-swings 20+20, (3 rounds)
  - squat & press 10+10, (2 rounds)
  - Push-ups (1 round)
  - shoulder mobility drill – prone with KB
- **Day 4**
  - x25 cleans each arm
- **Day 5 -** (14/2/20)
  - Unloaded TGU with foam pad (prop)
  - Deadlift (ladder to find max comfortable weight for >1 rep)
  - Snatch practice from - drop & catch
  - Deadlift (max reps to 7/10 RPE at max weight with 2 bells)
  - x10 2H swings, (2 rounds)

#### Week 2

- **Day 1** (17/2/20)
  - Unloaded TGU
  - Snatch practice (high pulls)
  - Goblet squat (3 rounds)
  - Swing, Clean, Press x10
  - Swing x10, Clean x10, Press x10
- **Day 2**
  - As many 8kg swings as possible
- **Day 3 -** (19/2/20)
  - 4-min carries
  - 2-handed swing drill with foam roller (example: http://ow.ly/ksoN30rKrWF)
  - Complex:
    - 10x 2H swing, 10+10 1H swing, Curl x10, Squat x10
    - 8x 2H swing, 8+8 1H swing, Curl x8, Squat x8
    - 6x 2H swing, 6+6 1H swing, Curl x6, Squat x8
    - 4x 2H swing, 4+4 1H swing, Curl x4, Squat x4
- **Day 4**
  - x20 single leg deadlifts (unloaded)
    https://www.youtube.com/watch?v=65WtVDlt4c8
- **Day 5 -** (21/2/20)
  - Bear crawls
  - Deadlift ladder
  - 100 swings: 5min
  - Press (5+5) + get-up

#### Week 3

- **Day 1 -** (24/2/20)
  - Swing, high-pull, snatch - 5+5+5 pause – 5 rounds (150 reps)
  - 2H swing + Deadlift - (25+20) (20+15) (15+10) – 1 min rest
  - 100 swings: 5min
- **Day 2**
  - Stand + Press from sitting, x20 each side https://youtu.be/-cW8HnKG09w

- **Day 3 -** (26/2/20)
  - Staggered stance cleans (10+10) x3
  - Press/push-press (5+5) x3
  - Suitcase march (2x 20 steps - slow)
- **Day 4**
  - x20 split squat/lunge – 4 available options from unloaded with support to loaded overhead without support.
  - Level 1 https://youtu.be/Cbd6PjV7rEk
  - Level 2 https://youtu.be/oBi8jJCFn8U
  - Level 3 https://youtu.be/ES3dLr28oRk
  - Level 4 https://youtu.be/hPScbrD0jFA
- **Day 5 -** (28/2/20)
  - 2 handed swings (to test heaviest appropriate weight)
  - Big 6 – set 1 (lunge of their choice in place of TGU)
    - 1 round - 2 rounds - 3 rounds - 2 rounds - 1 round
  - Big 6 – set 2
    - 2 reps - 3 reps

**Week 4**

- **Day 1 -** (02/3/20)
  - Complex (5 rounds)
    - Deadlift - 10, 8, 6, 4, 2 at approx. 60% 1RM
    - 1-arm row – 2, 3, 4, 5, 6 reps
    - Goblet squat – 2, 4, 6, 8, 10 reps
    - Press – 10, 8, 6, 4, 2 reps
    - 2H swings 10, 10, 10, 10, 10 reps
- **Day 2**
  - Bottom-up (3 options – standing, with side-step, with turn)
  - Option 1: https://youtu.be/O98zfiCm6lM
  - Option 2: https://youtu.be/Mz7eIGiJKKY
  - Option 3: https://youtu.be/1aKtr6qVIyg

- **Day 3 -** (04/3/20)
  - Complex (3 rounds, option for 4)
    - Seesaw press – 6, 8, 10 reps
    - Double front (box) squat – 6, 8, 10 reps
    - Single arm swing – 20, 20, 20 reps
    - Deadlift ladder – 3, 4, 5 reps on each (20 – max)
- **Day 4**
  Push-ups
- **Day 5 -** (06/3/20)
  - Snatch practice only

**Week 5**

- **Day 1 -** (09/3/20)
  - 2-handed swing ladder (1-10-1)
  - Snatch test – prep (60)
  - Goblet squat ladder
  - One-arm long cycle – 30/30
- **Day 2**
  - Split squat options for modifications provided: target 30 each side
  - Option 1: http://ow.ly/g6fJ30qohGi
  - Option 2: http://ow.ly/k5X530qohGz
- **Day 3 -** (11/3/20)
  - Max 5 rounds – in pairs at their pace
    - 1-arm row (5+5)
    - Goblet squat (5)
    - 1H swing (5+5)
    - Snatch (5+5)

      - Deadlift (5)
- Day 4
  - Goblet squat to chair (max) https://youtu.be/IloPf9Htz-E
- **Day 3 -** (13/3/20)
  - 100 swings: 5 mins
  - PBs (their choice of exercise), *with target reps*
    - 1H swing (5+5)
    - 2H swing (x10)
    - Clean (5+5)
    - Snatch (5+5)
    - Press (5+5)
    - Push-ups (as many as possible, if possible)
    - Bottom-up (able to balance only (no reps) on the strongest arm)
    - Goblet squat (x5) – *must be able to lift weight themselves*
    - Goblet squat (x25)

**Week 6**

- **Day 1 -** (16/3/20)
  - 2-handed swing (sideways) (4x 10)
  - Complex – 5 rounds (1-2-3-2-1)
    - Swing
    - High pull
    - Clean
    - Squat
    - Press
    - Half lunge/split squat (rear-step), left and right
- Day 2
  - Split squat options for modifications provided: target 30 each side
    - Option 1: Push-press https://youtu.be/0A8mapbpnZE
    - Option 2: Strict press https://youtu.be/kprCrEoNY0E
    - Option 3: Snatch https://youtu.be/NM_BwlPLV9s
- **Day 3 -** (18/3/20)
  - 100 swings: 5 mins - *with prescribed weight*
  - PBs – *with prescribed weight and some prescribed reps*
    - 1H swing (5+5)
    - 2H swing (x10)
    - Clean (5+5)
    - Snatch (5+5)
    - Press (5+5)
    - Push-ups (as many as possible, if possible)
    - Bottom-up (able to balance only (no reps) on the strongest arm)
    - Goblet squat (x5) – *must be able to lift weight themselves*
    - Goblet squat (x25)
    - TGU *(where possible)*
- Day 4 (19/3/20)
  - Loaded single leg deadlift
    - Option 1: With support: http://ow.ly/q1yT50yOLgl
    - Option 2: Without support: http://ow.ly/vt3F50yOMgs

**HOME TRAINING COMMENCED** (abbreviated test session at BIHS due to COVID-19)

- **Day 5 -** (20/3/20)
  - Complex – choice of 5 or 10 reps of each
    - 1. Staggered stance single arm deadlift
    - 2. Single arm swing
    - 3. Press (or push-press)
    - 4. Snatch

#### Week 7

- **Day 1 -** (23/3/20)
  - 1. 10x deadlifts
    2. 10x 2-handed swings
    3. 5+5 one-arm rows
    4. 10x goblet squats
    http://ow.ly/z4rd50yW548

  - Optional changes
    DEADLIFTS
    1. Staggered stance
    2. Single arm
    3. Change reps

    SWINGS
    1. 1-handed swings
    2. Change reps

    ROWS
    1. Change reps

    GOBLET SQUATS
    1. Change depth (use seat)
    2. Two bells

- **Day 2** (24/3/20)
  - The 'Any ol' get-up': http://ow.ly/vweH50yW3yc
    - In as many different ways as possible (no target set).
- **Day 3 -** (25/3/20)
  - 20x deadlift + 1x floor transfer
    15 seconds rest
  - 20x swings + 1x floor transfer
    15 seconds rest
  - Repeat (with 30s, 45s, 60s rest *if necessary*) until RPE = 8.
  - Options:
    If the weight isn't heavy enough, do 1-handed deadlifts and swings
- **Day 4** (26/3/20)
  - Arm bar
    - Until the shoulder starts to fatigue
- **Day 5 -** (27/3/20)
  - 300 swings
    - 300 swings a day challenge – their choice of swing accumulated throughout the day

#### Week 8

- **Day 1 -** (30/3/20)
  - 'Kettlebell shuffle': http://ow.ly/8ZgT50z1J5v
    - 55 or 91 reps of deadlift, 2-handed swing, goblet squat and press
- **Day 2** (31/3/20)
  - Shoulder depression in long-sitting: https://youtu.be/4K3SACf8q1I
    - Target: Accumulate 3 minutes throughout the day
- **Day 3 -** (1/4/20)
  - Hang-snatch to thruster: http://ow.ly/l4da30qu8hI
    - Target: As many repetitions as possible in *30 minutes MAX*
- **Day 4** (2/4/20)
  - Bear crawl: http://ow.ly/K1iS50z2EBE
    - Target: accumulate 3 minutes of moving bear crawls in any direction.
- **Day 5 -** (3/4/20)
  - 'Kettlebell shuffle': http://ow.ly/8ZgT50z1J5v
    - 55 or 91 reps of deadlift, 2-handed swing, goblet squat and press

#### Week 9

- **Day 1 -** (6/4/20) http://ow.ly/hE5z50z5Dat
    - Complex (1/5): Swing + Snatch + Press + Squat
        - Max (RPE 8) on non-dominant arm ▪ rest fully ▪ repeat on dominant arm
- **Day 2** (7/4/20)
    - Complex (2/5): Swing + Snatch + Press + Squat
        - Target: 1 more round each side with *no rest between arms*
- **Day 3 -** (8/4/20)
    - Complex (3/5): Swing + Snatch + Press + Squat
        - *Switch arms every 2 rounds.*
        - Target: 2 more rounds (combined total)
- **Day 4** (9/4/20)
    - Complex (4/5): Swing + Snatch + Press + Squat
        - *Switch arms every round*.
        - Target: 2 more rounds (combined total)
- **Day 5 -** (10/4/20)
    - Complex (5/5): Swing + Snatch + Press + Squat
        - As many as possible! Switch at will

#### Week 10

- **Day 1 -** (13/4/20)
    - 40spm swing cadence: http://ow.ly/IWKu50zcjm8
        - Start with an 8kg 2-handed, progressing with swing type (1 vs 2 hands, towel and weight as able) only once 40spm cadence has been achieved and comfortable.
- **Day 2** (14/4/20)
    - Loaded set-up – one set on each leg to failure: http://ow.ly/IWKu50zcjm8
        - Participant's choice of weight(s) and carry option: single or double suitcase, single or double rack carry, single or double overhead carry, asymmetrical carry
- **Day 3 -** (15/4/20)
    - Turkish Get-up: http://ow.ly/6zBL50zeig6
        - Practicing one or all transitions, with or without weight, depending on capability for up to 30 minutes.
- **Day 4** (16/4/20)
    - Loaded single leg deadlift using a wall as a guide, either sliding the rear foot along the floor or elevated, depending on capability – one set on each leg to failure: http://ow.ly/73b650zewp3
        - *S*
- **Day 5 -** (17/4/20)
    - Skills selection – participant's choice of 9 exercises:
        - Alternating single-arm swing

#### Week 11

- **Day 1 -** (20/4/20)
    - Complex of: row, swing, high-pull, snatch (option 2 only), clean & squat. Increasing by 1 rep each side with each round, as many rounds as possible
        - Option 1: http://ow.ly/Gowf30qyXYG
        - Option 2: http://ow.ly/T2il30qyXYL
- **Day 2** (21/4/20)
    - Shoulder taps (high plank or on knees): http://ow.ly/qlqU50ziHM4
        - As many as possible in one set
- **Day 3 -** (22/4/20)
    - Complex: *7 or 9* (reps) - http://ow.ly/IDR550zk6E8
        1. Single-leg deadlift
        2. One-arm row

3. Dead clean (i.e. from the floor)
4. Kickstand swing x5 then clean
5. Forward lunge (same side)
6. Backward lunge (same side)
7. Squat
8. Strict press (no leg drive)
9. Snatch

- **Day 4** (23/4/20)
  - Kick through: http://ow.ly/FuQA50zl4Yb
    - Target – 20 each side
- **Day 5 -** (24/4/20)
  - Complex – '*buns of steel*': http://ow.ly/KxHr50zm6YM
    - Single leg deadlift
    - Row
    - Dead clean
    - Split squat
    - Strict Press (optional)

**Week 12**

- **Day 1 -** (27/4/20)
  - **Volume load target** (Within group range: 30-60% more than the average for the previous 5 weeks)
- **Day 2** (28/4/20)
  - **Push-ups**; every minute on the minute. Technique: their choice of hands vs knees, Time: their choice of 10 minutes or 5 minutes. Target reps: their choice of 5 or 10 (or whatever they can manage).
- **Day 3 -** (29/4/20)
  - **Volume load target** (calculated as an approximate mid-way between Monday and Friday's values).
- **Day 4** (30/4/20)
  - **Table top** isometric hold
    - *For as long as possible.*
- **Day 5 -** (01/5/20)
  - **Volume load target** (Within group range: 10-290kg higher than current max load for the trial)

**Additional reference videos provided:**

| | | |
|---|---|---|
| Goblet squat (heels elevated): | | http://ow.ly/iIPr50yNopp |
| Clean: | | http://ow.ly/wbzu30qh62V |
| Clean (mid-pull): | | http://ow.ly/t0eS30qh63z |
| Clean (staggered stance - wall assisted): | | http://ow.ly/1LzM30qh64b |
| Hand assisted clean: | | http://ow.ly/NXjc30qh62B |
| Clean: | | http://ow.ly/gHiF30qieaU |
| Clean: | | http://ow.ly/8Iot30qieb6 |
| Deadlift: | | http://ow.ly/tQYP30qiebj |
| Snatch: | | http://ow.ly/2o0d30qiebp |
| Snatch: | | http://ow.ly/bmzg30qiecw |
| Snatch: | | http://ow.ly/EoPt30qiecP |
| Get-up: | | http://ow.ly/p9dO30qied2 |
| Two-handed swing: | | http://ow.ly/Z3gV30qiebS |
| Two-handed swing: | | http://ow.ly/gWXo30qiedb |
| Barbell assisted push-ups: | | http://ow.ly/5g1X30qoxOs |
| KB sport examples | (women): | http://ow.ly/eGjU50yNozk |
| | (men): | http://ow.ly/Oyhh50yNoEA |
| Clean & jerk | | http://ow.ly/7Zay50yNozR |
