## Supplementary material for "Effects of supervised high-intensity hardstyle kettlebell training on grip strength and health-related physical fitness in insufficiently active older adults: The BELL pragmatic controlled trial": Schedule of data collection

**Table 1.** Schedule of data collection

| Time point | Admission | Baseline | CONTROL | | TRAINING | | Follow-up |
|---|---|---|---|---|---|---|---|
| | Pre-trial | Test 1 (wk 0) | Test 2 (wk 4) | Test 3 (wk 13) | Test 4 (wk 19) | weeks 14-25 | Test 5 (wk 29) |
| **Enrollment** | | | | | | | |
| Eligibility screen | ● | | | | | | |
| Informed consent | ● | | | | | | |
| **Intervention** | | | | | | | |
| Kettlebell training | | | | | ● | ● | |
| **Primary outcome** | | | | | | | |
| Grip strength | | ● | ● | ● | ● | | ●*# |
| **Secondary outcomes** | | | | | | | |
| *Anthropometrics* | | | | | | | |
| Height & weight | | ● | ● | ● | | | ●* |
| *Cardiorespiratory endurance* | | | | | | | |
| 6-minute walk distance | | ● | ● | ● | | | ●*# |
| Stair climb time | | ● | ● | ● | ● | | ●* |
| Resting blood pressure | | ● | ● | ● | | | ●* |
| *Muscular strength and power* | | | | | | | |
| Knee extension strength | | ● | ● | ● | | | ●* |
| Hip extension strength | | ● | ● | ● | | | ●* |
| Sit to stand (app) | | ● | ● | ● | | | ●* |
| Counter movement vertical jump | | ● | ● | ● | ● | | ●* |
| *Muscular endurance* | | | | | | | |
| 30-second sit-to-stand | | ● | ● | ● | | | ●* |
| *Flexibility* | | | | | | | |
| Fingertip to floor | | ● | ● | ● | | | ●* |
| *Body composition* | | | | | | | |
| Dual-energy X-ray absorptiometry | | ● | ● | ● | | | ●* |
| Bioelectrical impedance | | ● | ● | ● | | | ●* |
| *Functional capacity* | | | | | | | |
| Five-times floor transfer | | ● | ● | ● | ● | | ●*# |
| 1 repetition maximum kettlebell deadlift | | ● | ● | ● | | | ●* |
| Static balance | | ● | ● | ● | | | ●* |
| *Quality of life* | | | | | | | |
| 36-item short form survey | | ● | ● | ● | ● | | ●*# |
| Sense of coherence | | ● | ● | ● | ● | | ●*# |
| *Training load* | | | | | | | |
| Training load volume (V-TL) | | | | | | ● | |
| Session rate of perceived exertion | | | | | | ● | |
| *Adherence, compliance, adverse events* | | | | | | ● | |
| *Interviews* | | | | | | | ● → |

*<70 years ($n$ = 11), # 70+ ($n$ = 15)
